# Neoantigen Landscape Supports Feasibility of Personalized Cancer Vaccine for Follicular Lymphoma

**DOI:** 10.1101/2022.01.06.22268805

**Authors:** Cody A. Ramirez, Felix Frenkel, Michelle Becker-Hapak, Erica K. Barnell, Ethan D. McClain, Sweta Desai, Timothy Schappe, Onyinyechi C. Onyeador, Olga Kudryashova, Vladislav Belousov, Alexander Bagaev, Elena Ocheredko, Susanna Kiwala, Jasreet Hundal, Zachary L. Skidmore, Marcus P. Watkins, Thomas B. Mooney, Jason R. Walker, Kilannin Krysiak, David A. Russler-Germain, Felicia Gomez, Catrina C. Fronick, Robert S. Fulton, Robert D. Schreiber, Neha Mehta-Shah, Amanda F. Cashen, Brad S. Kahl, Ravshan Ataullakhanov, Nancy L. Bartlett, Malachi Griffith, Obi L. Griffith, Todd A. Fehniger

## Abstract

Personalized cancer vaccines designed to target neoantigens represent a promising new treatment paradigm in oncology. In contrast to classical idiotype vaccines, we hypothesized that ‘polyvalent’ vaccines could be engineered for the personalized treatment of follicular lymphoma (FL) using neoantigen discovery by combined whole exome sequencing (WES) and RNA sequencing (RNA-Seq). Fifty-eight tumor samples from 57 patients with FL underwent WES and RNA-Seq. Somatic and B-cell clonotype neoantigens were predicted and filtered to identify high-quality neoantigens. B-cell clonality was determined by alignment of B-cell receptor (BCR) CDR3 regions from RNA-Seq data, grouping at the protein level, and comparison to the BCR repertoire of RNA-Seq data from healthy individuals. An average of 52 somatic mutations per patient (range: 2-172) were identified, and two or more (median: 15) high-quality neoantigens were predicted for 56 of 58 samples. The predicted neoantigen peptides were composed of missense mutations (76%), indels (9%), gene fusions (3%), and BCR sequences (11%). Building off of these preclinical analyses, we initiated a pilot clinical trial using personalized neoantigen vaccination combined with PD-1 blockade in patients with relapsed or refractory FL (#NCT03121677). Synthetic long peptide (SLP) vaccines were successfully synthesized for and administered to all four patients enrolled to date. Initial results demonstrate feasibility, safety, and potential immunologic and clinical responses. Our study suggests that a genomics-driven personalized cancer vaccine strategy is feasible for patients with FL, and this may overcome prior challenges in the field.

## Introduction

Follicular lymphoma (FL) is the most common indolent non-Hodgkin’s lymphoma,^1,2^ and remains predominantly incurable with conventional therapies.^3^ The clinical course of FL is heterogenous, whereby some patients experience minimal annual progression over decades,^4^ whereas others rapidly progress, or transform into aggressive diffuse large B-cell lymphoma (DLBCL). In an attempt to identify less toxic alternatives to traditional chemotherapy-based approaches, the anti-CD20 monoclonal antibody (mAb) rituximab was approved as the first immunotherapy for FL patients, and demonstrated improvements in progression-free and overall survival in clinical trials.^5–7^ Even with combination therapy of rituximab and chemotherapy, ∼35-45% of patients with advanced disease relapse within five-years of initial treatment.^8^ While more recent advances include the use of the immunomodulatory agent lenalidomide, various targeted small molecule inhibitors, and chimeric antigen receptor (CAR) T-cells, there remains a need to develop novel therapies to optimally balance response rate, duration of treatment and responses, as well as short-and long-term toxicities.^9,10^

Personalized cancer vaccines stimulate a patient’s own immune system to specifically attack cancer cells. Such vaccines target neoantigens, which are short mutated peptide sequences that are specifically expressed by tumor cells. Major histocompatibility complex (MHC) class I and/or II molecules have demonstrated the ability to present neoantigens to cytotoxic and/or helper T-cells for recognition and antitumor immune responses.^11–14^ Reports of neoantigen vaccination showed promise in various solid cancers,^15–31^ and CD8+ T-cell responses were identified against 10 driver mutations in a retrospective cohort of FL patients.^32^ Prior efforts to develop cancer vaccines for FL had the notable weaknesses of targeting only a single lymphoma target (the dominant B-cell receptor (BCR) idiotype), and were not tested in conjunction with potentially synergistic immune checkpoint inhibitors, which may have contributed to the negative results from previous phase III clinical trials.^33–35^ We hypothesized that a combined polyvalent approach that collectively identifies multiple cancer vaccine targets (BCR clonotypes, small somatic mutations, and gene fusions) could aid in personalized immunotherapy for patients with FL.

Defining the feasibility of neoantigen vaccines, optimizing effective vaccine design informed by next-generation sequencing data, and identifying factors that contribute to the success of personalized cancer vaccines are important areas of inquiry.^36,37^ FL typically has a low-to-medium mutation burden,^38,39^ and in a report that evaluated 117 FL tumors,^40^ the median number of somatic variants per individual was 55 (range: 2-169). However, the proportion of somatic FL-specific mutations that create high-quality predicted neoantigens is not well described. Therefore, given that the correlation between mutation burden and immunotherapy response is modest, and that even low numbers of highly immunogenic neoantigens can precipitate a detectable response, testing for neoantigens within FL is supported.^32^ Here we report on neoantigen vaccine design in FL using a comprehensive genomic approach (**Figure 1**). We applied whole exome (WES) and RNA sequencing (RNA-Seq) to a retrospective cohort of 58 samples from 57 patients with FL, and then performed neoantigen prediction and *in silico* vaccine design. This strategy was then employed on 4 novel relapsed or refractory (rel/ref) FL patients to assess efficacy of treatment with personalized neoantigen vaccine therapy in combination with PD-1 blockade (#NCT03121677).

**Figure 1:**
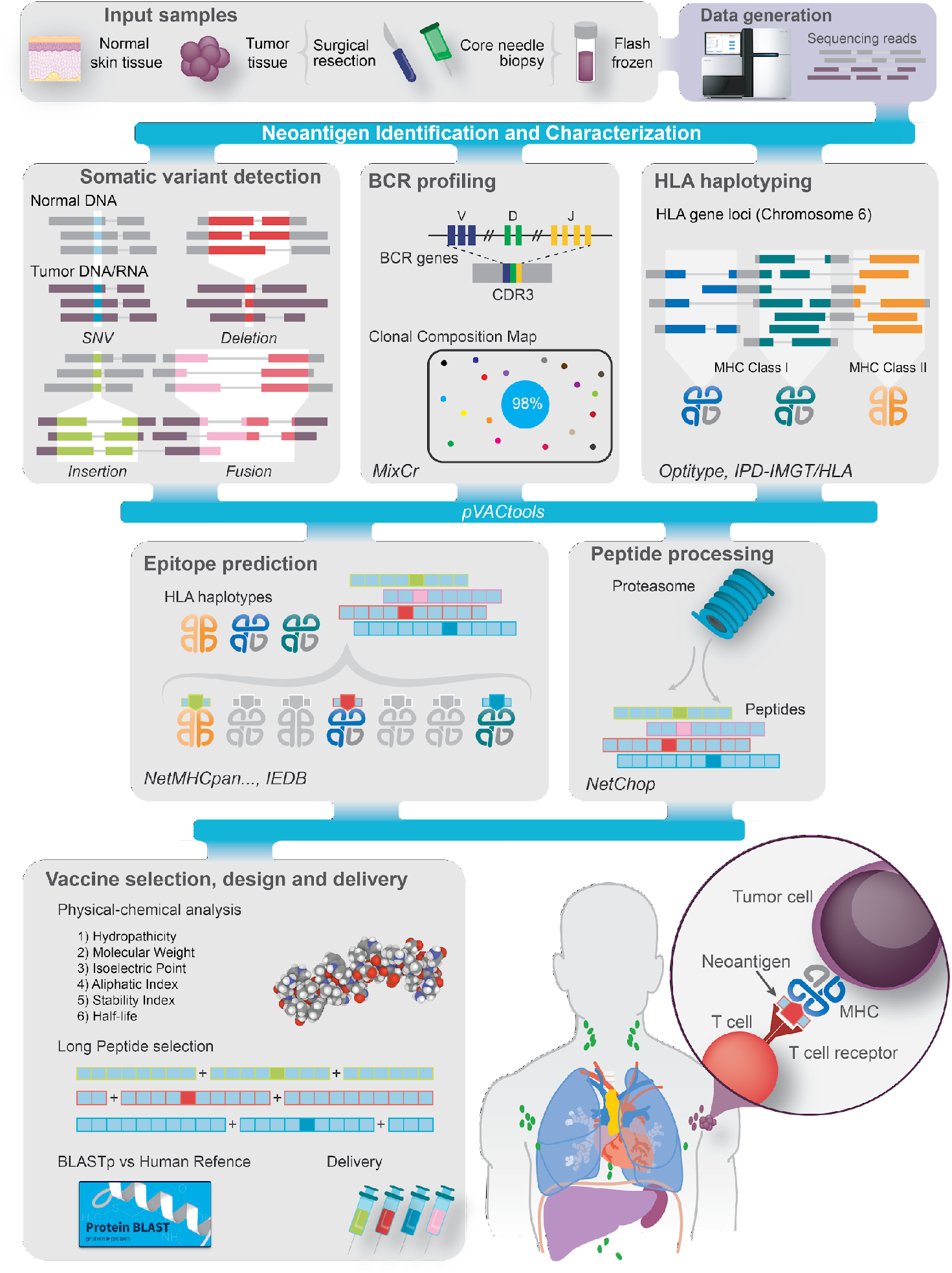
Overview of the FL personalized cancer vaccine pipeline. Patient samples are acquired and then sequenced (top left). Somatic variants of various types, including single nucleotide variants (SNVs; *blue*), deletions (*red*), insertions (*green*), and fusions (*pink*), are predicted. Sequence data are analyzed to determine human leukocyte antigen (HLA) types and B-cell clonotypes for each patient. Variant and clonal B-cell peptide sequences are inferred from variants and analyzed with respect to their predicted expression, proteasome processing, and ability to bind the patient’s MHC Class I complexes. Candidates are then selected for vaccine design and additional analyses are performed to assess manufacturability. Bioinformatic tools used for each step are indicated in *italics. Abbreviations*: *BCR*, B-cell receptor; *CDR3*, complementarity-determining region 3; *IEDB*, Immune Epitope Database.

## Results

### Whole exome/transcriptome sequencing reveals the diverse mutational landscape of FL

To predict high-quality neoantigen candidates for FL personalized cancer vaccines, we performed WES on 58 retrospectively collected fresh-frozen tumor samples with paired non-malignant tissue from 57 unique patients. RNA-Seq was performed on 57 of the 58 tumor samples. In total, our cohort included 27 patients with treatment-naive FL, 21 with relapsed disease, eight with transformed FL, and one with composite lymphoma (**Table S1**). WES for all samples (tumor and normal) achieved >20X coverage for >75% of the targeted region, with a mean coverage of 76X. RNA-Seq for all samples averaged 145 million total reads (range: 48-545 million) with an average of 67% of reads mapped (range: 36-98%). For the RNA-Seq data, the average breakdown of aligned bases was: 6.3% ribosomal (range: ∼0-45%), 11% UTR (range: ∼9-27%), 6% intronic (range: ∼1-42%), 1% intergenic (range: ∼0-7%) and 76% coding (range: ∼14-87%). After filtering, the number of nonsynonymous coding variants per sample ranged from 2-172 variants (mean: 52; median: 36). Of the 1,787 affected genes, 264 were mutated in more than one patient. Thirty-two of 39 genes that were previously identified as significantly mutated by Krysiak et al.^40^ were also found to be mutated within our cohort. For our cohort, many genes with established relevance to FL were recurrently mutated (*KMT2D*/MLL2 [67%], *CREBBP* [41%], *TNFRSF14* [41%], *BCL2* [36%], *ATP6V1B2* [16%], *STAT6* [12%], *EZH2* [10%], *IRF8* [10%], *CD79B* [10%], *BCL7A* [9%], *EP300* [9%], *MEF2B* [9%], *CARD11* [7%], *TP53* [7%], and *GNA13* [7%]; **Figure 2, Table S2**). Many individuals harbored more than one *KMT2D* mutation (57 mutations observed in 39 individuals) and/or more than one *BCL2* mutation (31 mutations observed in 21 individuals). No novel hotspot mutations were identified. Additionally, 40% (23 of 57 patients) harbored unique fusion genes (**Table S3; Figure S1**).

**Figure 2:**
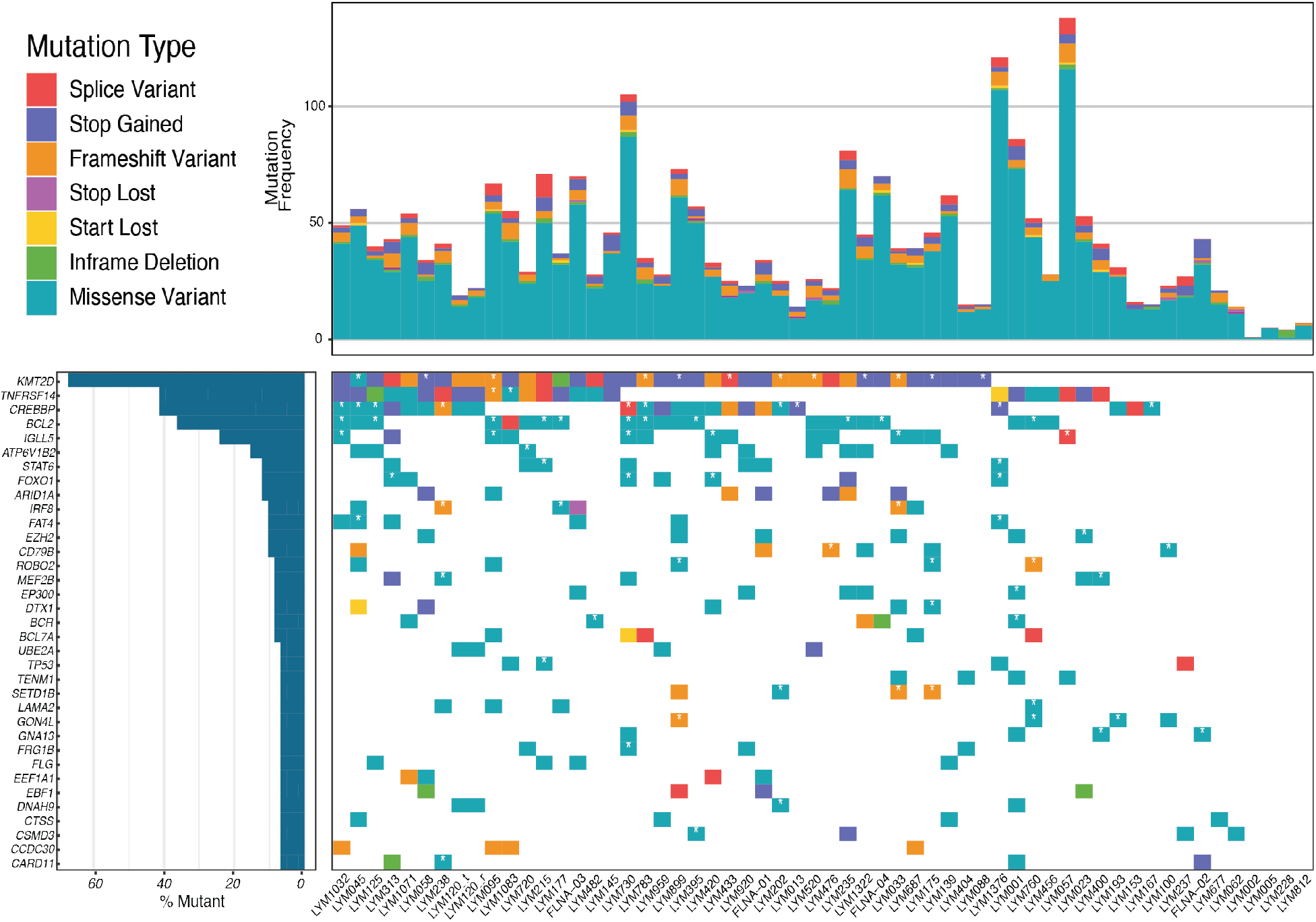
Recurrently mutated genes and mutation burden observed for all patients. The bar graph on the top corresponds to the number of total mutations per patient and is colored by mutation type. The bar graph on the left corresponds to the percentage of mutations for a given gene for the entire cohort. Columns represent each sample in the cohort (one patient, LYM120, with both tumor (t) and relapse (r) samples is shown) and are ordered by the presence of mutations in the most to least frequently mutated gene. The third plot indicates the presence or absence of a mutation for each patient and gene combination, colored by mutation type. If a patient has multiple mutations for an individual gene, it is colored according to the priority order as indicated in the mutation type legend, from top to bottom. A white star indicates which mutations are predicted to result in high quality neoantigen vaccine candidates.

### BCR clonality analysis of normal B-cells and FL identifies oligoclonal and polyclonal populations

To investigate B-cell clonal composition in normal lymph nodes and FL, we compared our tumor RNA-Seq data to publicly available RNA-Seq data of B-cell enriched samples.^41–49^ In total, 53 normal and 57 tumor samples had sufficient BCR sequencing coverage for analysis. Comparison of the reconstructed BCR repertoire revealed that normal samples showed diverse polyclonality (median IgH clonality = 0.04) whereas malignant FL samples showed low polyclonality (median IgH clonality = 0.25) (**Figure 3; Figure S2**). To establish a malignant-specific metric, we used a dominant clonotype group threshold of 9%. Among the 53 normal samples, only two (3.8%) had a dominant IgH clonotype exceeding the selected cut-off, whereas most FL samples (78%) had a likely malignant clonotype group exceeding the 9% cut-off. Analysis of clonotype composition for light chains (IgL and IgK) revealed a similar pattern.

**Figure 3:**
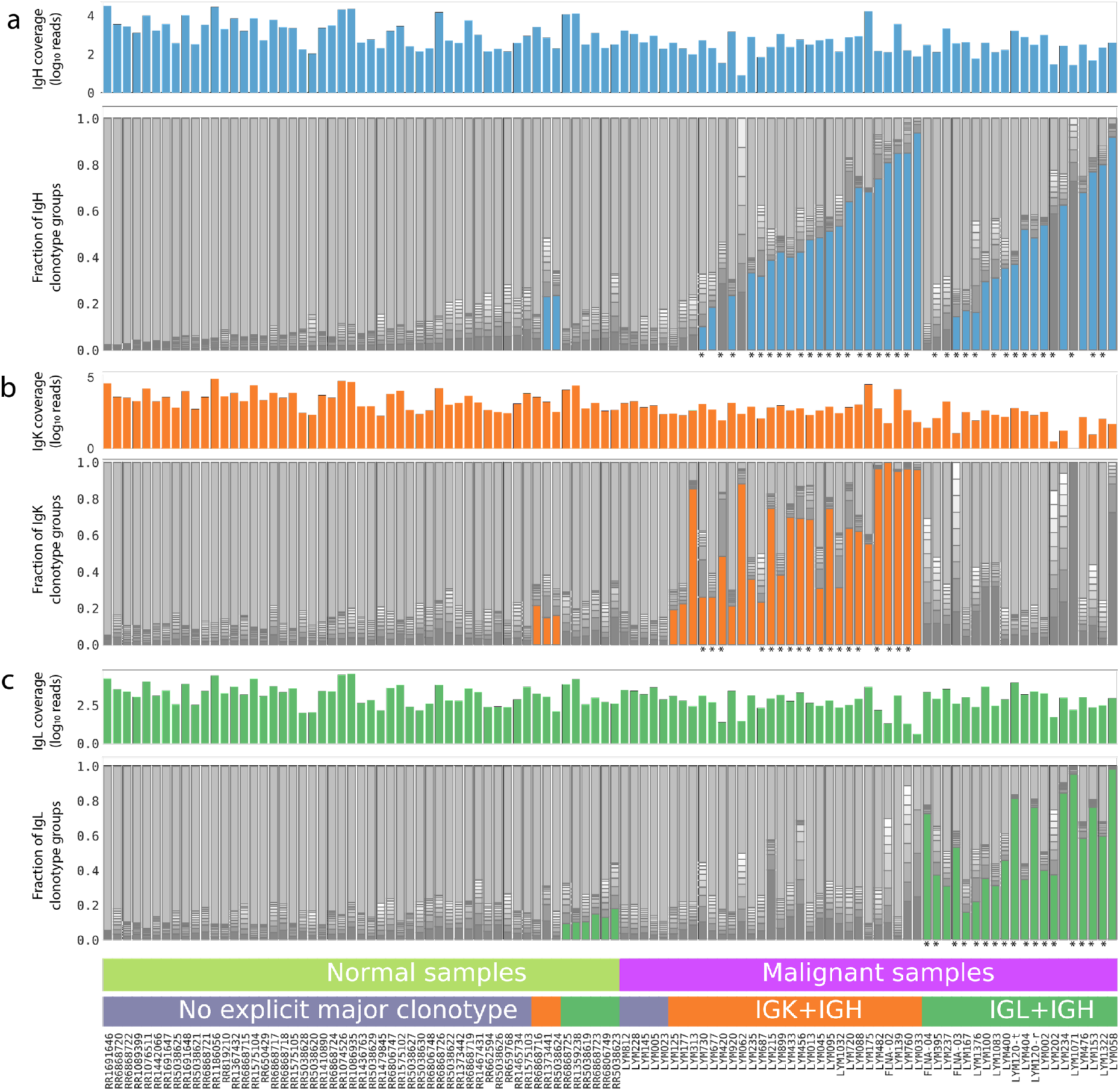
Clonality analysis of B-cell receptor populations within healthy normal samples and follicular lymphoma samples. The plots below show the composition of BCR repertoire of both normal (left half) and malignant (right half) samples for each of Ig chains separately: panels a, b and c correspond to heavy (blue), kappa (orange) and lambda (green) chains respectively. In each panel the upper histogram shows the coverage of the given Ig chain in the sample (log_10_ of read counts) while the lower stacked barplot shows BCR repertoire structure. Each bar from bottom to top is composed of ten dark sections representing the fraction of repertoire for the ten largest clones in the sample and a single top light gray section representing all other (minor) clones. The colored bottom sections depict the single most dominant clone that exceeded the cutoff value of fraction in the sample (9%) and had sufficient overall coverage (>40 reads for malignant samples while normal samples were preselected having >100 reads for each chain, see Methods). When both light chains passed the cutoff only the one with the larger fraction was selected and colored. Stars below each panel indicate major clonotypes predicted to result in one or more high quality neoantigen vaccine candidates.

Given that FLs express a unique cell surface BCR and selectively retain expression of the BCR for survival^50^, FL neoantigen candidates can be derived from the immunoglobulin heavy (IgH) and/or light (lambda/kappa, IgL/K) chains in addition to somatic variants (SNVs/indels). The average number of total clonotypes (IgH, IgL, and IgK) detected within each patient’s BCR repertoire was 1,400 (median: 550, range: 20-22,052). An average of 361 unique IgH clonotypes (range: 6-7,762) and an average of 770 unique IgL/K clonotypes (range: 6-10,029) were identified. A total of 59 out of 20,244 (0.3%) unique IgH clonotypes and 57 out of 43,918 (0.1%) unique IgL/K clonotypes were found to be major or minor clones. A total of 116 unique clonal (i.e., >9%) BCRs (mode: 2, range: 0-6) were identified (**Figure 3, Figure S3a**,**b**,**f**). An average of 5,083 reads (range: 136-76,742) were used in clonotyping each patient’s BCR repertoire (**Figure S3c**). The average number of independent supporting reads per clonotype was four (range: 2-13). Of the 57 patients with RNA-Seq data, 46 had at least one clonal IgH and IgL/K clonotype, two patients had clonal IgL/K clonotypes only, one patient had clonal IgH clonotypes only, and eight had no clonal IgH or IgL/K clonotype.

Interestingly, 17 patients had two or more dominant IgH and/or IgL/K clones. To determine if multiple dominant clones displayed subclonal architecture, a pairwise nucleotide and protein sequence alignment between all dominant clones within a patient’s BCR repertoire was assessed (**Figure S3g**,**h; Table S4**,**S5**). Dominant clones with the same VDJ alleles had greater sequence similarities when compared to dominant clones with different VDJ alleles. For example, patient LYM720 had four dominant IgH clones at 25%, 24%, 17%, and 13%. Three of the clones shared the exact same VDJ alleles based on the best protein alignment score percentage to each other (range of 93-98%) (**Table S4**). However, when these three dominant clones were compared to the fourth clone with a differing VDJ allele, the best alignment score percentage was 49%. This finding, in conjunction with somatic variant allele frequency (VAF) from WES data, implies that the LYM720’s clone 1 is the founding FL clone while clones 2 and 3 represent subclones. Clone 4 shares little sequence similarity to the other clones and may represent an independant malignant clone or B-cell expansion from an unrelated immune response. A similar pattern was observed for patients with multiple IgL/K dominant clones (**Table S5**). After manual review of the IgL/K clonotype pairwise protein sequence alignment data, we concluded that five patients have multiple dominant clonotypes with no relation to each other while another five patients have a founding and subclonal relationship as previously described (e.g., see patient LYM120; **Figure S3g**). The dominant IgL/K clones also displayed high CDR3 nucleotide sequence homology to each other (1 nucleotide change). We hypothesize that Ig somatic hypermutation likely accounts for the majority of changes in BCR clonality observed within our cohort (**Figure S3h**).

### High-quality personalized neoantigen cancer vaccine candidates were predicted for most patients

Using the WashU analysis pipeline (pVACtools)^51^, 3,065 high-quality somatic variants were identified (**Figure 4a**). Of these, 783 were predicted to be small, high-quality somatic variants with neoantigen potential (**Figure 4a; Table S6**). Peptides suitable for cancer vaccine generation were identified for all but two patients from our cohort and 55/57 (96%) patients had at least two vaccine candidates (**Figure 4b,c**). These predicted high-quality neoantigens were associated with known driver variants as well as presumed passenger mutations (**Figure S4**). For the 42 fusion variants identified across all patients, 25 gene fusions (median: 2, range: 0-4) were predicted to generate high-quality neoantigen vaccine candidates (**Figure S1, Table S3**). These fusions were identified in 25% (14/57) of our FL cohort (**Figure 4c**,**S5; Table S7**). In total, 22% (26/116) of the BCR clonotypes identified as dominant clones included short peptide sequences predicted to sufficiently bind to their respective MHCs (500nm < IC50 binding affinity ≤ 1,000nm), while 61% (71/116) included short peptide sequences that are particularly strong predicted binders to their respective MHCs (IC50 binding affinity ≤ 500nm). A total of 97 clonal BCRs (mode: 2, range: 0-6) were predicted to be high-quality neoantigen vaccine candidates, representing 79% (45/57) of our FL cohort (**Figure 4c, S3f; Table S8**). Two or more neoantigen candidates were identified for 95% (55/58) samples with a mean of 16 predicted neoantigens per patient (range: 0-38). Overall, 76% (702/905) of the total predicted patient peptides arose from missense mutations, 9% (81/905) from indels, 3% (25/905) from gene fusions and 11% (97/905) from clonal BCR sequences. Furthermore, 97% (56/58) of samples had at least one missense or indel somatic vaccine candidate, 25% (14/57) of patients had at least one fusion vaccine candidate, 79% (45/57) of patients had at least one BCR vaccine candidate, and only one patient had no vaccine candidates. When compared to an orthogonal approach for neoantigen prediction (BostonGene Vaccine Module), our approach yielded more predicted neoantigen candidates, which is likely attributable to use of a less conservative requirement surrounding RNA-Seq support (**Table S9**,**10**). Twenty-one percent of vaccine candidates predicted were shared out of the total 1,179 collectively predicted vaccine candidates from the two approaches. For >76% of samples, a set of common neoantigens was generated by both approaches (**Figure S6**).

**Figure 4:**
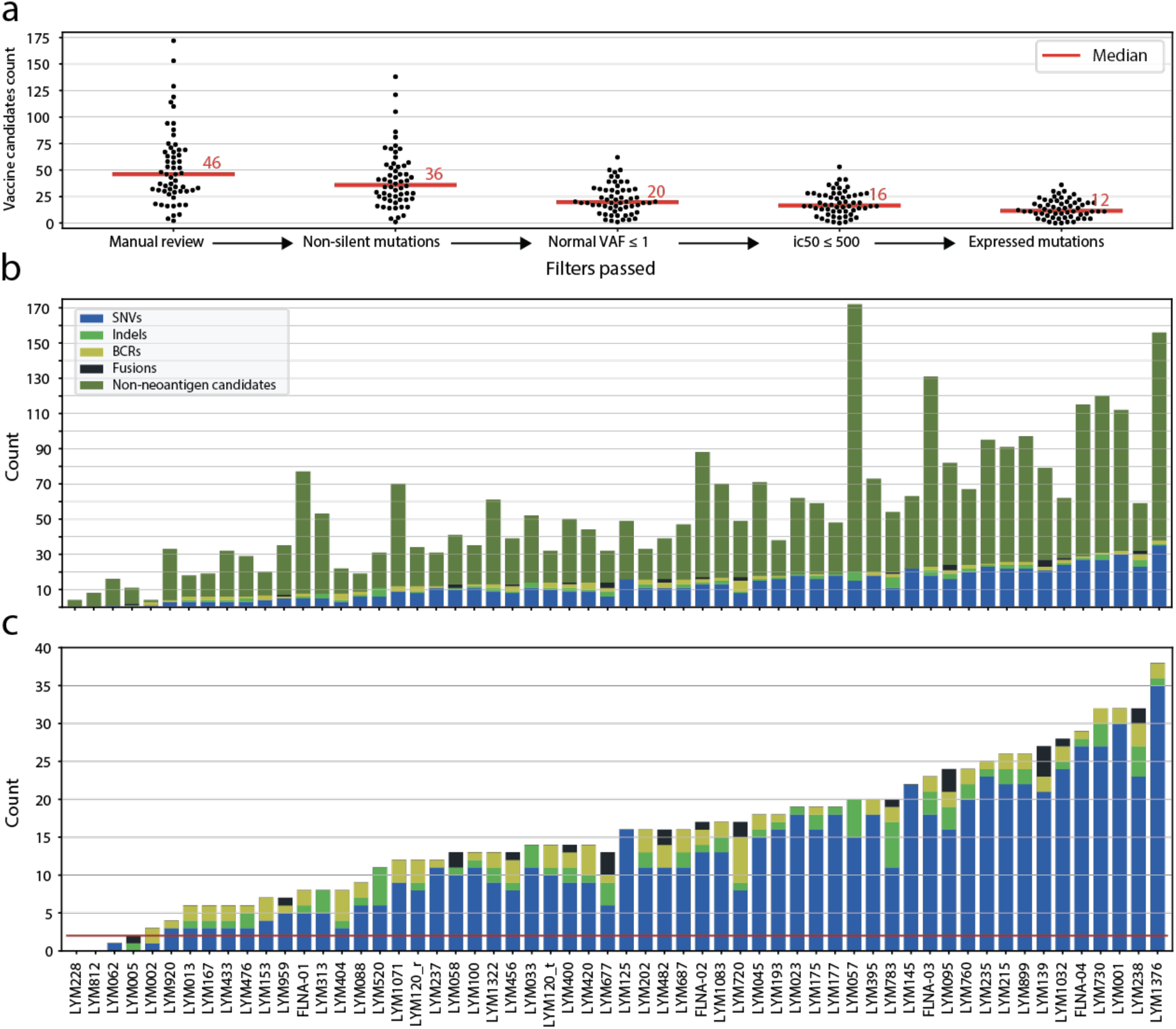
Personalized neoantigen cancer vaccine identification and prioritization. (a) Swarm plots display the number of vaccine candidates on the y-axis for the entire cohort at each stage of filtering (x-axis), moving from left to right. The bar graphs depict the numbers of final neoantigen vaccine candidates for each patient, colored according to source (SNV, indel, BCR, fusion) and are sorted from least to most total candidates. Bar graph (b) includes non-neoantigen mutations that did not pass neoantigen filtering, while bar graph (c) only contains final candidates. The red line in panel (c) depicts the minimum cutoff of 2 candidates required for potential vaccine design.

### Antigen presenting machinery (APM) assessment

Neoantigens are recognized by antigen-specific T-cells via presentation by MHC on the tumor cell surface. When investigating mutations in APM genes, we identified two samples bearing likely loss-of-function mutations (**Figure 5a**). A novel frameshift insertion in the β2 microglobulin (*B2M*) gene was observed in the beginning of the transcript (ENST00000558401:p.Ala6ArgfsTer52) for sample LYM1376. Another likely loss-of-function (nonsense) variant was found in the tapasin (*TAPBP*) gene (ENST00000426633:p.Gln477Ter) in LYM730. When investigating mutations in genes associated with MHC, no nonsynonymous mutations were identified; however, some samples had low expression of class I genes (e.g. LYM395, LYM760, and LYM1071), while others had low expression of class II genes (e.g. LYM228, LYM1376, and LYM235) (**Figure 5b**). Similar patterns have been previously described.^52^

**Figure 5:**
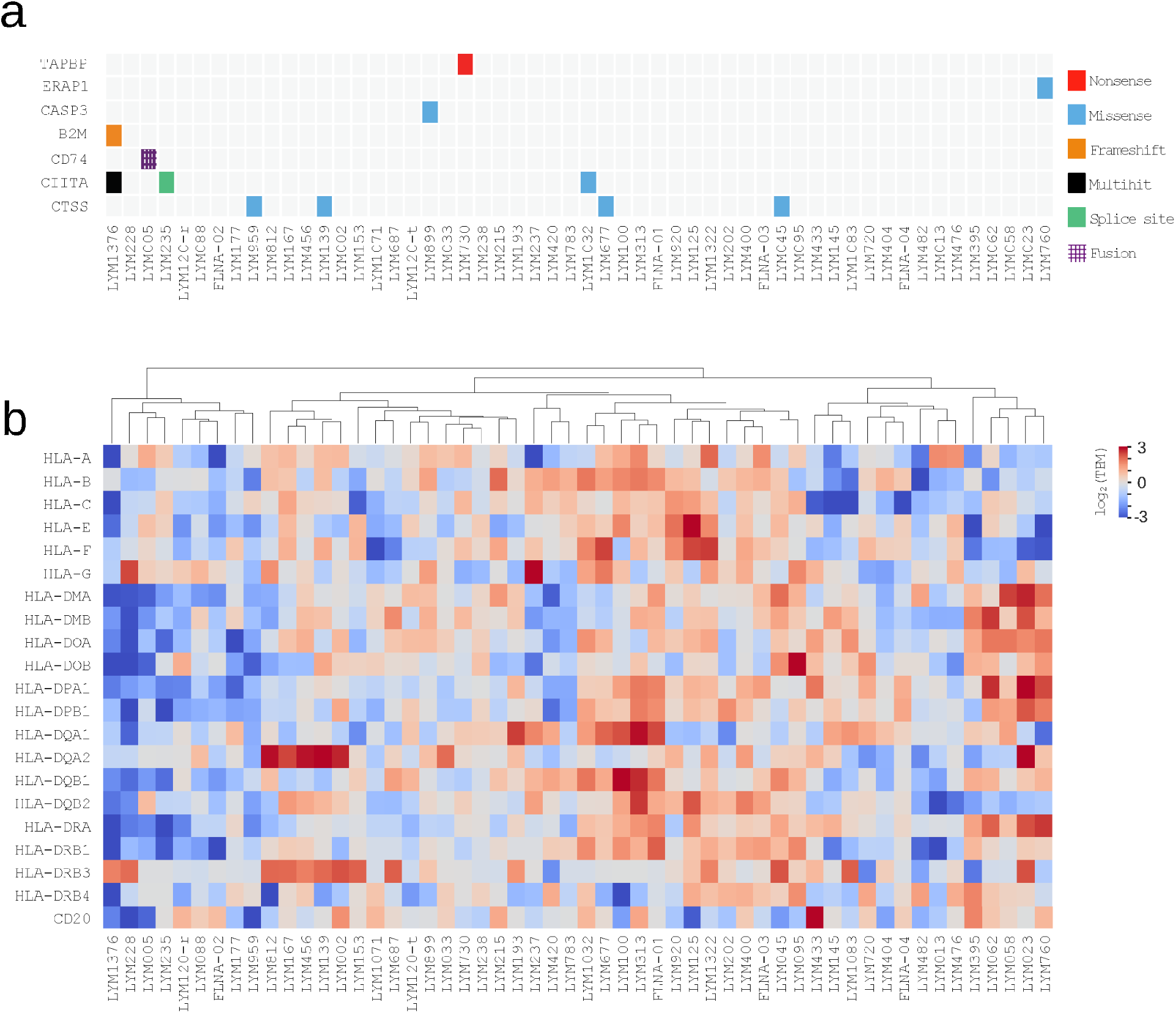
Molecular functional profiling of antigen presentation machinery and MHC. (a) Somatic mutations in antigen presenting machinery (APM) genes. Only 7 mutated genes of the total 32 APM genes considered. (b) Depicts median transformed log_2_(TPM+1) RNA expression levels of MHC Class I and II genes. *Abbreviations:* ssGSEA = single sample gene set enrichment analysis

### FL clonal architecture informs vaccine design

We investigated the impact of clonal architecture on vaccine design for 58 FL tumor samples. The average variant read depth per patient was 173 (range: 60-554) (**Figure S7**). The average VAF per patient was 23% (range: 7.6-40.9%) (**Figure S8**). The average number of tumor (sub)clonal populations identified per patient was 2 (range: 1-4) (**Figure S9**). Eleven (20%) patients were considered monoclonal whereas 43 (80%) were considered oligoclonal. The average tumor DNA VAF for predicted high-quality neoantigens was 26.3% (range: 5.0-97.7%). Most patients (94%) had at least one predicted high-quality neoantigen in the dominant/founding clone (median: 11; range: 0-30). Most patients (72%) with evidence of subclonality also had evidence of subclonal neoantigens. For example, patient LYM1376, with the most predicted high-quality neoantigens (36 small somatic mutations), had five and 25 high-quality neoantigens predicted within all clonal and subclonal populations, respectively (**Figure S9; Table S6**). Of note, the clonality analysis utilized only SNVs/indels as input, which underestimates the number of high-quality neoantigens observed.

### A pilot clinical trial shows preliminary safety and efficacy of a neoantigen vaccine plus anti-PD-1 mAb therapy in rel/ref FL

A pilot clinical trial (#NCT03121677) was initiated using personalized neoantigen vaccine therapy combined with PD-1 blockade in patients with rel/ref FL (**Figure S10**,**S11; Table S13**). To date, four patients have been enrolled. Pretreatment biopsies were used to identify 12-19 neoantigens for each patient. Peptide vaccines were synthesized using the predicted neoantigens and administered concurrently with nivolumab, an anti-PD-1 monoclonal antibody. Time from biopsy to treatment ranged from five to seven months. There were rare grade 1-2 adverse events (e.g., injection site reaction), and no grade 3-5 adverse events. Responses assessed by positron emission tomography (PET) and/or computerized tomography (CT) scanning after cycle 2 (C2) included one complete response (CR), one stable disease (SD), and two instances of progressive disease (PD). The two patients with PD after C2 received four weekly doses of rituximab, per protocol, in combination with additional neoantigen vaccine and nivolumab doses. Of these two patients, one achieved a partial response (PR) and one achieved a CR. Both continued on study with vaccine, nivolumab, and four additional doses of rituximab.

Correlative studies from the patient with CR post-C2 of vaccine plus nivolumab revealed a neoantigen-specific immune response (**Figure 6a**). For this patient, we identified 19 predicted high-quality neoantigen vaccine candidates using pVACtools (**Figure 6b**). A total of 16 peptides were formulated into a SLP vaccine, of which 13 were predicted to bind to HLA-A*68:01 and five were predicted to bind to HLA-A*23:01. After completing therapy, nine unique HLA-A*68:01 candidate peptides were screened with a newly created TAP-negative cell line to assess if neoantigens stabilized cognate MHC class I molecules. Four neoantigen candidates stabilized HLA-A*68:01 in a dose-dependent manner (3 of 4 positives shown, HIST1H2BK ENSP00000349430.1:p.A111X, ZNF100 ENSP00000445201.3:p.I218V, and BCL2 ENSP00000329623.3:p.A4T, **Figure 6c**). One of the candidates (CTIF ENSP00000256413.3:p.D495G) that did not stabilize HLA-A*68:01 on the TAP-negative cell line was selected as a non-binding control for subsequent assays. The long vaccine peptides were used to stimulate and expand peripheral blood mononuclear cells (PBMC) taken from the patient four months after initiation of the vaccine study therapy (**Figure 6d,e**). The greatest IFN-gamma response by ELISpot was observed for HIST1H2BK A111X (1,282 +/-30 SPU)/1e6 PBMCs) while the other two candidates ZNF100 I218V and BCL2 A4T showed 573+/-35, and 566+/-70 SPU/1e6 PBMCs, respectively. Both of the latter candidate SPUs were above the non-binding control peptide CTIF D459G 278+/-50 SPU/1e6, indicating a positive IFN-gamma response. To determine if circulating antigen-specific CD8+ T-cells could be detected in post-vaccination PMBCs, MHC class I tetramers were prepared with the three candidate peptides shown to be enriched by ELISPOT assay and used to probe ten-day, SLP-stimulated PBMCs. We found 0.5% of the CD8+ T cells were specific for HIST1H2BK A111X PE/APC tetramers, which was greater than the CTIF D459G non-binding control (**Figure 6f**). Lastly, ten-day post-vaccination PBMC cultures were stimulated with HIST1H2BK A111X SLP, and then re-stimulated with artificial antigen presenting cell (AAPC) loaded with candidate SLP. IFN-gamma/TNF expression from CD8+ T-cells was comparable to the positive control (**Figure 6g**). The same cultures, pulsed with the CTIF D459G non-binding control peptide, demonstrated a low cytokine response. Together, these results indicate that tumor-specific antigens can be used to stimulate antigen-specific T-cells for a patient with FL.

**Figure 6:**
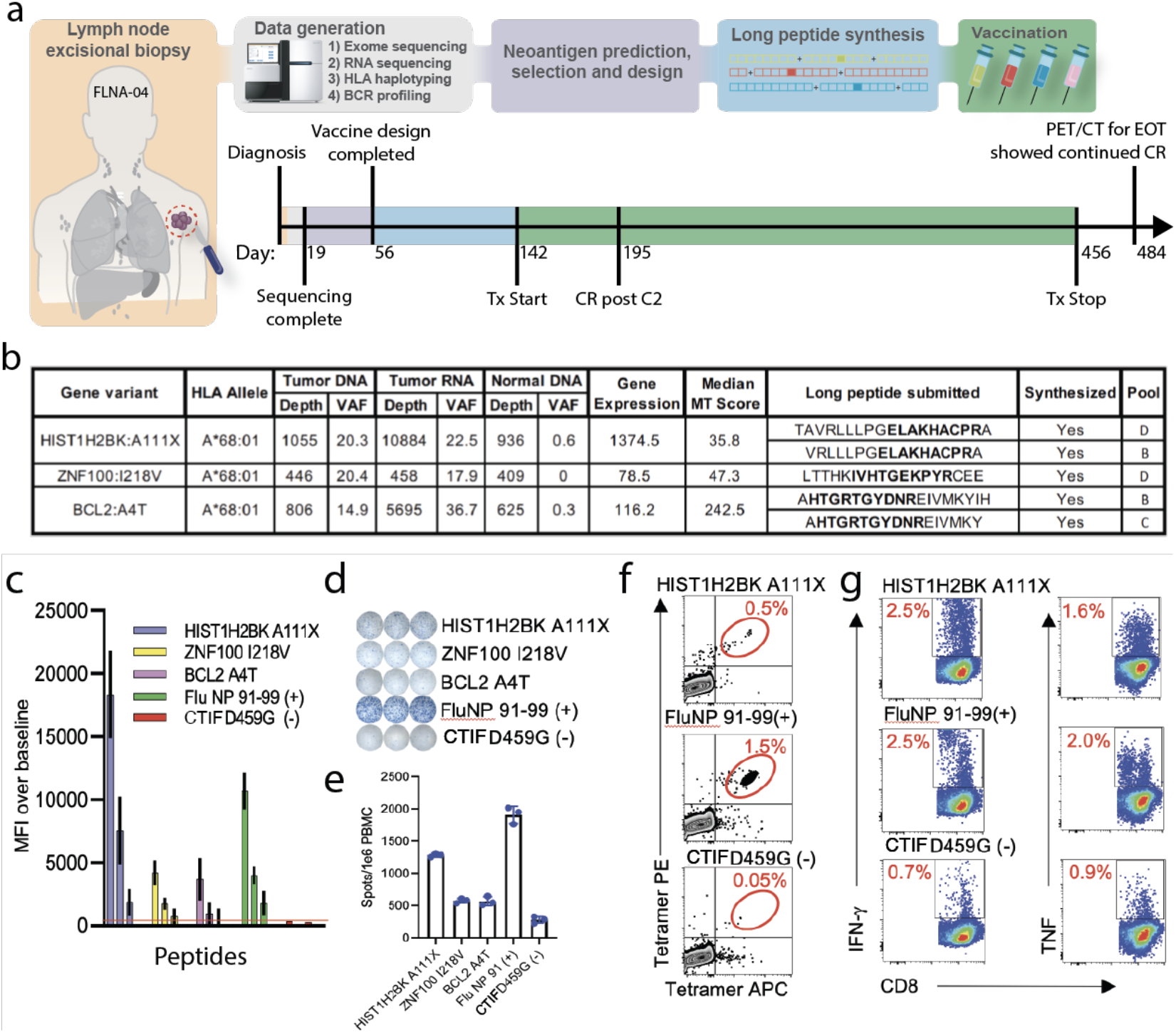
Implementation of neoantigen vaccines in pilot clinical trial. (a) Trial overview and timeline for patient FLNA-04 (See Suppl Figure 11 and 12 for more detailed versions). (b) The table lists all variants that were predicted to result in high quality neoantigen vaccine candidates along with corresponding HLA allele, selection criteria, long peptide sequence submitted, synthesis success status, and vaccination pool (**Table S11**,**S12**). Short epitope sequences that binding predictions were based on are bolded within the long peptide sequences submitted column. (c) Peptide stabilization of selected candidate neoantigens. Various concentrations of peptides were incubated overnight with ICP-47 expressing (TAP deficient) B cell line expressing HLA A*68:01 heavy chain, washed and then stained with W6/32 APC. Mean fluorescence intensity (MFI) of cells pulsed with decreasing peptide concentrations were compared to no peptide pulsed control cells to validate predicted peptide stabilization of MHC class I molecule. (d) PBMC from cycle 6 apheresis were pulsed with vaccinating peptides and cultured for twelve days in vitro and then challenged with predicted short peptides overnight on IFN-g coated ELISPOT plates. (e) Bar graph shows triplicate value spots per million PBMC from D12 culture. (f) An example positive antigen specific CTL enrichment detected by peptide loaded tetramers stained with Tetramer PE and Tetramer APC gated on live, CD3/CD8DP CTL from D12 CTL. (g) Antigen specific CTL from D12 cultures were challenged with artificial antigen presenting cells pulsed with specific peptides, incubated for six hours and then IFN-g or TNF expressing CD3/CD8 DP cells were detected by FACS. *Abbreviations: Tx = Treatment*, CR = Complete remission, C2 = Cycle 2, PET/CT = Positron emission tomography-computed tomography, EOT = End of Treatment, PBMC = peripheral blood mononuclear cell, VAF = variant allele frequency, MT = mutant, PE = phycoerythrin, APC = Antigen-presenting cells.

## Discussion

In this study, we performed comprehensive exome and transcriptome profiling to define the feasibility of using a neoantigen vaccine approach for the treatment of patients with FL. This included HLA typing, mutation calling, BCR clonotyping, fusion calling, neoantigen prediction, assessment of mutations in APM/MHC complexes, and sequencing-based *in silico* vaccine design for 58 FL samples from 57 patients. Our analyses indicated that the modest mutational burden of FL does not preclude patients from harboring immunogenic neoantigens that could be used for a personalized vaccine strategy. We further demonstrated that ‘polyvalent’ vaccine design is possible using a broad genomics-driven strategy. In this study, 97% of patients had at least one peptide suitable for cancer vaccine development, and 95% of patients had multiple vaccine candidates. Our approach identified high-quality neoantigens that were associated with genes implicated in FL, suggesting that known driver variants (as well as passenger variants) may play important roles in vaccine design. This preclinical proof-of-principle effort was translated into an on-going clinical trial to examine the safety and preliminary efficacy of neoantigen vaccines for patients with relapsed FL. This clinical trial was further corroborated by correlative studies of patient T-cell responses.

Previous attempts to develop personalized tumor vaccines for patients with FL used a strategy that targeted lymphoma-specific immunoglobulins (i.e., BCRs). These approaches utilized a single cancer target and patients often received cytotoxic chemotherapy in the interim, which likely contributed to key clinical trials failing to meet their primary efficacy endpoints.^33–35^ Emerging evidence indicates that targeting *multiple* neoantigens improves immune elimination of a tumor^12^ and recent studies have increased the number of neoantigens incorporated into cancer vaccines.^16,17,53,54^ These ‘polyvalent’ personalized neoantigen cancer vaccines may enhance the anti-tumor immune response and improve clinical efficacy of cancer vaccines.^55^

In this study, cancer vaccine design was informed by FL’s unique mutational landscape. In total, 67% of our cohort contains at least one predicted high-quality neoantigen vaccine candidate within a previously identified set of recurrently mutated genes in FL.^40,56–59^ For example, 25% of our cohort contained at least one *KMT2D* (*MLL2*) predicted high-quality neoantigen vaccine candidate. *KMT2D* mutations are present in ∼50% of FLs^40^ and play a clear role in FL pathogenesis, synergizing with *BCL2* overexpression in mouse models.^60^ They have also been identified as some of the earliest mutations acquired after the canonical *IgH-BCL2* t(14;18) initiating translocation event,^60^ which makes *KMT2D* neoantigens particularly attractive for FL cancer vaccines, since they are typically associated with the founding clone.^60,61^ Additionally, 40% (23/57) of the cohort harbored unique fusion genes (**Figure S1; Table S3**). The most recurrently identified fusion gene pair was a frameshift *ARL17A*--*KANSL1*, which has been implicated in other disease types, but not in FL.^62–64^ However, this lesion has also been described in normal cohorts,^65^ and its significance remains uncertain. *BCL6* was also involved in fusions with different gene partners occurring within four patients. A total of 25 gene fusions were predicted to be high-quality neoantigen vaccine candidates within 25% of the entire cohort (**Figure 4c**). Overall, somatic point mutations and fusions in known FL driver genes contributed significantly to vaccine design.

While known driver genes were regularly featured in our vaccine designs, recurrently mutated driver hotspots were not predicted to be high-quality neoantigens, which implies that FL vaccine designs will require patient-specific customization. Of the 783 predicted high-quality neoantigens resulting from small somatic mutations, only six were shared between two patients and only one was shared between three patients. Furthermore, due to differences in HLA alleles between individuals, even a shared variant does not imply a shared neoantigen. For example, of the six variants shared between two patients, only three were predicted to be high-quality neoantigens when accounting for patient HLA alleles. Similarly, the BCL2 variant R129H shared by three different patients (LYM783, LYM235, and LYM730) all resulted in different neoantigen candidates (TPFTARG-H-F, FTARG-H-FATV, and H-FATVVEEL) due to each patient’s specific HLA alleles. These observations suggest that patient-specific vaccines will be needed to improve patient outcomes compared to a universal approach.

Further, personalized cancer vaccines must consider both clonal and subclonal tumor populations. Tumors are heterogeneous and frequently evolve, creating subclones that are spatially and temporally separated.^66–70^ Vaccines targeting clonal neoantigens should induce a comprehensive anti-tumor response to eliminate the entire tumor population. However, targeting subclonal-specific neoantigens may protect against escape mechanisms (immune editing) where a subclone has selectively lost immunogenic neoantigens present in the founding clone.^71,72^ We show preliminary data suggesting that high-quality neoantigens can be identified in both clonal and subclonal tumor cell populations for most FL patients.

Several limitations and future work remain to be addressed to improve personalized cancer vaccines for FL. It is currently unclear what factors result in the ultimate success or failure of personalized cancer vaccines,^36,37^ and clinical trial results from other cancers published to date show a low accuracy for neoantigen prediction pipelines.^73^ Factors that could influence vaccine response include: 1) neoantigen identification pipelines, 2) HLA typing predictions, expression, and mutations, 3) peptide processing prediction, 4) MHC binding predictions, 5) vaccine design/delivery approach including neoantigen vaccine form, 6) T-cell recognition, and 7) tumor microenvironment. Existing tools either require further optimization, or have yet to be incorporated into our pipeline.

Our retrospective analysis suggests that nearly all FL patients may be candidates for personalized cancer vaccine clinical trials. Recent studies support polyvalent neoantigen cancer vaccines combined with an anti-PD-1 mAb to stimulate neoantigen-specific T-cells that were generated by the vaccine.^16,54^ These preclinical results led to our ongoing, first-in-human personalized neoantigen vaccine trial, that uses up to 20 SLP neoantigens in combination with anti-PD-1 mAb in patients with rel/ref FL (#NCT03121677). Preliminary results are encouraging with no serious adverse events and with CR observed in one of four evaluable patients to date. These pilot clinical trials are the ultimate test of bioinformatic predictions and will permit improvement of future neoantigen prediction pipelines. This study supports ongoing early phase clinical trial assessment of neoantigen vaccines in lymphoma, which match the goals of a chemotherapy-free immunotherapy without serious adverse events.

## Methods

Fresh-frozen tumor samples with paired non-malignant tissue (skin or blood) were retrospectively collected. All patients provided written informed consent for the use of their samples in sequencing and all clinical characteristics are summarized in **Table S1**. Samples with known FL status underwent WES and RNA-Seq. Somatic and B-cell clonotype neoantigens were predicted and filtered to identify high-quality neoantigens. B-cell clonality was determined by alignment of B-cell receptor (BCR) CDR3 regions from RNA-Seq data, grouping at the protein level, and comparison to the BCR repertoire of RNA-seq data from healthy individuals. The analysis pipeline considered all small somatic mutations (SNVs and indels), gene fusions, and BCR dominant clones that passed filtering criteria and manual inspection as neoantigen candidates. Neoantigen candidates were considered for both clonal and subclonal populations. Subsequently, a pilot clinical trial was initiated using personalized neoantigen vaccination combined with PD-1 blockade in patients with relapsed FL (#NCT03121677). Pretreatment biopsies were used to identify multiple unique high-quality tumor-specific neoantigens (12-19 neoantigens per patient) for all enrolled patients. SLP vaccines were successfully synthesized for all four patients, and for each patient, ∼20 peptides (including alternate registers for the same neoantigen, as needed) were pooled into four groups of five, to minimize competition for the same MHC molecule. Each of the four pools were administered to patients via subcutaneous injection into different limbs with concurrent intravenous nivolumab administration. See **Supplementary Methods** for additional details.

## Data Availability

All data have been deposited in dbGaP (accession: phs001229).

## Acknowledgements

We thank the patients and their families for the donation of their samples and participation in clinical trials. This work was supported by the Michael and Ena Feinberg Lymphoma Research Fund, the Jamie Erin Follicular Lymphoma Research Consortium, the Steinback fund of the Barnes Jewish Hospital Foundation, and the Alvin J. Siteman Cancer Center Siteman Investment Program (supported by The Foundation for Barnes-Jewish Hospital, Cancer Frontier Fund and Barnard Trust). We also thank the Siteman Cancer Center at Washington University School of Medicine for the use of the Siteman Flow Cytometry Core, which provided cell sorting service and the Bursky Center for Human Immunology and Immunotherapy Immunomonitoring Laboratory. The Siteman Cancer Center is supported in part by an NCI Cancer Center Support Grant P30CA091842. Obi Griffith was supported by the National Cancer Institute (NCI) of the National Institutes of Health (NIH) on awards U01CA209936, U01CA231844, U24CA237719, and U01CA248235. Malachi Griffith was supported by the National Human Genome Research Institute (NHGRI) of the NIH under award R00HG007940, the NCI under awards U01CA209936, U01CA231844, U24CA237719 and U01CA248235, and the V Foundation for Cancer Research under award V2018-007. We would also like to acknowledge that **Figure 1** is adapted and modified from **Figure 1** of Richters et al. 2019. Genome Medicine. 11(1):56.

## Authorship Contributions

R.D.S., N.M.S., A.F.C., N.L.B., B.S.K., R.A., M.G., O.L.G., and T.A.F. conceptualized the study. C.A.R., F.F., M.B.H., O.K., R.D.S., N.L.B., M.G., O.L.G., and T.A.F. developed the experimental design. C.A.R., M.B.H., E.D.M., S.D., T.S., and M.P.W. acquired samples and clinical data. T.B.M, J.R.W., C.C.F., and R.S.F. led sequence library construction and data acquisition. C.A.R., F.F., M.B.H., E.D.M., S.D., O.K., V.B., A.B., E.O., and Z.L.S. performed data analysis. S.K., J.H., V.B., E.O., F.F., and J.R.W. developed software for neoantigen analysis. M.B.H., E.D.M., S.D., T.S., and O.C.O. performed immunologic response studies. C.A.R., F.F., O.K., M.B.H., and O.L.G. prepared figures and tables. C.A.R., F.F., M.B.H., O.K., E.K.B, and O.L.G. wrote the manuscript with input from K.K., F.G., D.A.R.G., M.G., and T.A.F. All authors approved the final version of the manuscript.

## References

1. Linet MS, Vajdic CM, Morton LM, et al. Medical history, lifestyle, family history, and occupational risk factors for follicular lymphoma: the InterLymph Non-Hodgkin Lymphoma Subtypes Project. J. Natl. Cancer Inst. Monogr. 2014;2014(48):26–40.

2. Project N-HLC. A clinical evaluation of the International Lymphoma Study Group classification of non-Hodgkin’s lymphoma. Blood, The Journal of the American Society of Hematology. 1997;89(11):3909–3918.

3. Kahl BS, Yang DT. Follicular lymphoma: evolving therapeutic strategies. Blood. 2016;127(17):2055–2063.

4. Ardeshna KM, Smith P, Norton A, et al. Long-term effect of a watch and wait policy versus immediate systemic treatment for asymptomatic advanced-stage non-Hodgkin lymphoma: a randomised controlled trial. Lancet. 2003;362(9383):516–522.

5. Schulz H, Bohlius JF, Trelle S, et al. Immunochemotherapy With Rituximab and Overall Survival in Patients With Indolent or Mantle Cell Lymphoma: A Systematic Review and Meta-analysis. J. Natl. Cancer Inst. 2007;99(9):706–714.

6. Hiddemann W, Kneba M, Dreyling M, et al. Frontline therapy with rituximab added to the combination of cyclophosphamide, doxorubicin, vincristine, and prednisone (CHOP) significantly improves the outcome for patients with advanced-stage follicular lymphoma compared with therapy with CHOP alone: results of a prospective randomized study of the German Low-Grade Lymphoma Study Group. Blood. 2005;106(12):3725–3732.

7. Maloney DG, Grillo-López AJ, Bodkin DJ, et al. IDEC-C2B8: results of a phase I multiple-dose trial in patients with relapsed non-Hodgkin’s lymphoma. J. Clin. Oncol. 1997;15(10):3266–3274.

8. Flinn IW, van der Jagt R, Kahl B, et al. First-Line Treatment of Patients With Indolent Non-Hodgkin Lymphoma or Mantle-Cell Lymphoma With Bendamustine Plus Rituximab Versus R-CHOP or R-CVP: Results of the BRIGHT 5-Year Follow-Up Study. J. Clin. Oncol. 2019;37(12):984–991.

9. Kahl B. High-risk follicular lymphoma: Treatment options. Hematol. Oncol. 2021;39 Suppl 1:94–99.

10. Jacobson CA, Chavez JC, Sehgal AR, et al. Axicabtagene ciloleucel in relapsed or refractory indolent non-Hodgkin lymphoma (ZUMA-5): a single-arm, multicentre, phase 2 trial. Lancet Oncol. 2022;23(1):91–103.

11. Schumacher TN, Schreiber RD. Neoantigens in cancer immunotherapy. Science. 2015;

12. Yarchoan M, Johnson BA III, Lutz ER. Targeting neoantigens to augment antitumour immunity. Nat. Rev. 2017;

13. Schumacher TN, Scheper W. Cancer neoantigens. Annual review of. 2019;

14. Chu Y, Liu Q, Wei J, Liu B. Personalized cancer neoantigen vaccines come of age. Theranostics. 2018;8(15):4238.

15. Robbins PF, Lu YC, El-Gamil M, Li YF, Gross C. Mining exomic sequencing data to identify mutated antigens recognized by adoptively transferred tumor-reactive T cells. Nat. Med. 2013;

16. Ott PA, Hu Z, Keskin DB, et al. An immunogenic personal neoantigen vaccine for patients with melanoma. Nature. 2017;

17. Keskin DB, Anandappa AJ, Sun J, Tirosh I. Neoantigen vaccine generates intratumoral T cell responses in phase Ib glioblastoma trial. Nature. 2019;

18. Tran E, Ahmadzadeh M, Lu Y-C, et al. Immunogenicity of somatic mutations in human gastrointestinal cancers. Science. 2015;350(6266):1387–1390.

19. Cohen CJ, Gartner JJ, Horovitz-Fried M, et al. Isolation of neoantigen-specific T cells from tumor and peripheral lymphocytes. J. Clin. Invest. 2015;125(10):3981–3991.

20. Gros A, Parkhurst MR, Tran E, et al. Prospective identification of neoantigen-specificlymphocytes in the peripheral blood of melanoma patients. Nat. Med. 2016;22(4):433–438.

21. Tran E, Turcotte S, Gros A, et al. Cancer immunotherapy based on mutation-specific CD4+ T cells in a patient with epithelial cancer. Science. 2014;344(6184):641–645.

22. Linnemann C, van Buuren MM, Bies L, et al. High-throughput epitope discovery reveals frequent recognition of neo-antigens by CD4+ T cells in human melanoma. Nat. Med. 2015;21(1):81–85.

23. Strønen E, Toebes M, Kelderman S, et al. Targeting of cancer neoantigens with donor-derived T cell receptor repertoires. Science. 2016;352(6291):1337–1341.

24. Wick DA, Webb JR, Nielsen JS, et al. Surveillance of the tumor mutanome by T cells during progression from primary to recurrent ovarian cancer. Clin. Cancer Res. 2014;20(5):1125– 1134.

25. Pritchard AL, Burel JG, Neller MA, et al. Exome Sequencing to Predict Neoantigens in Melanoma. Cancer Immunol Res. 2015;3(9):992–998.

26. Koşaloğlu Z, Zörnig I, Halama N, et al. Identification of immunotherapeutic targets by genomic profiling of rectal NET metastases. Oncoimmunology. 2016;5(11):e1213931.

27. Anagnostou V, Smith KN, Forde PM, et al. Evolution of Neoantigen Landscape during Immune Checkpoint Blockade in Non-Small Cell Lung Cancer. Cancer Discov. 2017;7(3):264–276.

28. Rizvi NA, Hellmann MD, Snyder A, et al. Mutational landscape determines sensitivity to PD-1 blockade in non–small cell lung cancer. Science. 2015;348(6230):124–128.

29. George S, Miao D, Demetri GD, et al. Loss of PTEN Is Associated with Resistance to Anti-PD-1 Checkpoint Blockade Therapy in Metastatic Uterine Leiomyosarcoma. Immunity. 2017;46(2):197–204.

30. Le DT, Durham JN, Smith KN, et al. Mismatch repair deficiency predicts response of solid tumors to PD-1 blockade. Science. 2017;357(6349):409–413.

31. Mennonna D, Maccalli C, Romano MC, et al. T cell neoepitope discovery in colorectal cancer by high throughput profiling of somatic mutations in expressed genes. Gut. 2017;66(3):454–463.

32. Nielsen JS, Sedgwick CG, Shahid A, et al. Toward Personalized Lymphoma Immunotherapy: Identification of Common Driver Mutations Recognized by Patient CD8+ T Cells. Clin. Cancer Res. 2016;22(9):2226–2236.

33. Schuster SJ, Neelapu SS, Gause BL, et al. Vaccination with patient-specific tumor-derived antigen in first remission improves disease-free survival in follicular lymphoma. J. Clin. Oncol. 2011;29(20):2787–2794.

34. Levy R, Ganjoo KN, Leonard JP, et al. Active idiotypic vaccination versus control immunotherapy for follicular lymphoma. J. Clin. Oncol. 2014;32(17):1797–1803.

35. Freedman A, Neelapu SS, Nichols C, et al. Placebo-controlled phase III trial of patient-specific immunotherapy with mitumprotimut-T and granulocyte-macrophage colony-stimulating factor after rituximab in patients with follicular lymphoma. J. Clin. Oncol. 2009;27(18):3036–3043.

36. Wells DK, van Buuren MM, Dang KK, et al. Key Parameters of Tumor Epitope Immunogenicity Revealed Through a Consortium Approach Improve Neoantigen Prediction. Cell. 2020;183(3):818–834.e13.

37. Richters MM, Xia H, Campbell KM, et al. Best practices for bioinformatic characterization of neoantigens for clinical utility. Genome Med. 2019;11(1):56.

38. Lawrence MS, Stojanov P, Polak P, et al. Mutational heterogeneity in cancer and the search for new cancer-associated genes. Nature. 2013;499(7457):214–218.

39. Alexandrov LB, Nik-Zainal S, Wedge DC, et al. Signatures of mutational processes in human cancer. Nature. 2013;500(7463):415–421.

40. Krysiak K, Gomez F, White BS, et al. Recurrent somatic mutations affecting B-cell receptor signaling pathway genes in follicular lymphoma. Blood. 2017;129(4):473–483.

41. Lonsdale J, Thomas J, Salvatore M, et al. The Genotype-Tissue Expression (GTEx) project. Nat. Genet. 2013;45(6):580–585.

42. Shchetynsky K, Diaz-Gallo L-M, Folkersen L, et al. Discovery of new candidate genes for rheumatoid arthritis through integration of genetic association data with expression pathway analysis. Arthritis Res. Ther. 2017;19(1):19.

43. Mo A, Marigorta UM, Arafat D, et al. Disease-specific regulation of gene expression in a comparative analysis of juvenile idiopathic arthritis and inflammatory bowel disease. Genome Med. 2018;10(1):48.

44. Dvinge H, Bradley RK. Widespread intron retention diversifies most cancer transcriptomes. Genome Med. 2015;7(1):45.

45. Chen R, Xia L, Tu K, et al. Longitudinal personal DNA methylome dynamics in a human with a chronic condition. Nat. Med. 2018;24(12):1930–1939.

46. Madan V, Kanojia D, Li J, et al. Aberrant splicing of U12-type introns is the hallmark of ZRSR2 mutant myelodysplastic syndrome. Nat. Commun. 2015;6:6042.

47. Dvinge H, Ries RE, Ilagan JO, et al. Sample processing obscures cancer-specific alterations in leukemic transcriptomes. Proc. Natl. Acad. Sci. U. S. A. 2014;111(47):16802– 16807.

48. Visconte V, Rogers HJ, Singh J, et al. SF3B1 haploinsufficiency leads to formation of ring sideroblasts in myelodysplastic syndromes. Blood. 2012;120(16):3173–3186.

49. Ferreiro JF, Rouhigharabaei L, Urbankova H, et al. Integrative Genomic and Transcriptomic Analysis Identified Candidate Genes Implicated in the Pathogenesis of Hepatosplenic T-Cell Lymphoma. PLoS ONE. 2014;9(7):e102977.

50. Küppers R. Mechanisms of B-cell lymphoma pathogenesis. Nat. Rev. Cancer. 2005;

51. Hundal J, Kiwala S, McMichael J, et al. pVACtools: A Computational Toolkit to Identify and Visualize Cancer Neoantigens. Cancer Immunol Res. 2020;8(3):409–420.

52. Green MR, Kihira S, Liu CL, et al. Mutations in early follicular lymphoma progenitors are associated with suppressed antigen presentation. Proc. Natl. Acad. Sci. U. S. A. 2015;112(10):E1116–25.

53. Carreno BM, Magrini V, Becker-Hapak M, et al. Cancer immunotherapy. A dendritic cell vaccine increases the breadth and diversity of melanoma neoantigen-specific T cells. Science. 2015;348(6236):803–808.

54. Sahin U, Derhovanessian E, Miller M, et al. Personalized RNA mutanome vaccines mobilize poly-specific therapeutic immunity against cancer. Nature. 2017;547(7662):222– 226.

55. Hellmann MD, Snyder A. Making It Personal: Neoantigen Vaccines in Metastatic Melanoma. Immunity. 2017;47(2):221–223.

56. Pasqualucci L, Khiabanian H, Fangazio M, et al. Genetics of follicular lymphoma transformation. Cell Rep. 2014;6(1):130–140.

57. Green MR, Gentles AJ, Nair RV, et al. Hierarchy in somatic mutations arising during genomic evolution and progression of follicular lymphoma. Blood. 2013;121(9):1604–1611.

58. Okosun J, Bödör C, Wang J, et al. Integrated genomic analysis identifies recurrent mutations and evolution patterns driving the initiation and progression of follicular lymphoma. Nat. Genet. 2014;46(2):176–181.

59. Correia C, Schneider PA, Dai H, et al. BCL2 mutations are associated with increased risk of transformation and shortened survival in follicular lymphoma. Blood. 2015;125(4):658–667.

60. Ortega-Molina A, Boss IW, Canela A, et al. The histone lysine methyltransferase KMT2D sustains a gene expression program that represses B cell lymphoma development. Nat. Med. 2015;21(10):1199–1208.

61. Zhang J, Dominguez-Sola D, Hussein S, et al. Disruption of KMT2D perturbs germinal center B cell development and promotes lymphomagenesis. Nat. Med. 2015;21(10):1190– 1198.

62. Zhou JX, Yang X, Ning S, et al. Identification of KANSARL as the first cancer predisposition fusion gene specific to the population of European ancestry origin. Oncotarget. 2017;8(31):50594–50607.

63. López-Nieva P, Fernández-Navarro P, Graña-Castro O, et al. Detection of novel fusion-transcripts by RNA-Seq in T-cell lymphoblastic lymphoma. Sci. Rep. 2019;9(1):5179.

64. Mehani B, Narta K, Paul D, et al. Fusion transcripts in normal human cortex increase with age and show distinct genomic features for single cells and tissues. Sci. Rep. 2020;10(1):1368.

65. Boettger LM, Handsaker RE, Zody MC, McCarroll SA. Structural haplotypes and recent evolution of the human 17q21.31 region. Nat. Genet. 2012;44(8):881–885.

66. Gerlinger M, Rowan AJ, Horswell S, et al. Intratumor Heterogeneity and Branched Evolution Revealed by Multiregion Sequencing. N. Engl. J. Med. 2012;366(10):883–892.

67. de Bruin EC, McGranahan N, Mitter R, et al. Spatial and temporal diversity in genomic instability processes defines lung cancer evolution. Science. 2014;346(6206):251–256.

68. McGranahan N, Furness AJS, Rosenthal R, et al. Clonal neoantigens elicit T cell immunoreactivity and sensitivity to immune checkpoint blockade. Science. 2016;351(6280):1463–1469.

69. Milo I, Bedora-Faure M, Garcia Z, et al. The immune system profoundly restricts intratumor genetic heterogeneity. Science Immunology. 2018;3(29.):

70. Gejman RS, Chang AY, Jones HF, et al. Rejection of immunogenic tumor clones is limited by clonal fraction. Elife. 2018;7:e41090.

71. Linette GP, Becker-Hapak M, Skidmore ZL, et al. Immunological ignorance is an enabling feature of the oligo-clonal T cell response to melanoma neoantigens. Proceedings of the National Academy of Sciences. 2019;116(47):23662–23670.

72. Schaettler MO, Richters MM, Wang AZ, et al. Characterization of the Genomic and Immunological Diversity of Malignant Brain Tumors Through Multi-Sector Analysis. Cancer Discovery. 2021;candisc.0291.2021.

73. Linette GP, Carreno BM. Neoantigen Vaccines Pass the Immunogenicity Test. Trends Mol. Med. 2017;23(10):869–871.

