## Supplemental Data for "Neoantigen Landscape Supports Feasibility of Personalized Cancer Vaccine for Follicular Lymphoma"

**List of Supplementary Data Items**

**Supplementary Methods**

**Supplementary Figure 1**: WashU manually reviewed fusion genes waterfall plot

**Supplementary Figure 2**: Comparative healthy and malignant B-cell repertoire

**Supplementary Figure 3**: B-cell tumor repertoire oligoclonality analysis identifies oligoclonal and polyclonal populations within follicular lymphoma patients

**Supplementary Figure 4:** Recurrently mutated genes and mutation burden observed for all predicted high quality neoantigens with patients

**Supplementary Figure 5**: WashU Predicted high quality fusion neoantigen vaccine candidates waterfall plot

**Supplementary Figure 6**: Predicted high quality neoantigen predictions and comparison between WashU and BostonGene analysis pipelines

**Supplementary Figure 7**: Clonality read depth

**Supplementary Figure 8**: Clonality DNA VAF

**Supplementary Figure 9**: Inferring subclonal architecture objectively in follicular lymphoma patients

**Supplementary Figure 10**: Clinical trial treatment schema (#NCT03121677)

**Supplementary Figure 11**: Overview of pilot trial to evaluate neoantigen vaccine + ICT in FL

**Supplementary Table 1:** Clinical characteristics of patients used for genetic analysis

#### **Supplementary Table 2:** VCF file containing WashU manually reviewed short somatic variants

**Supplementary Table 3**: WashU manually reviewed gene fusion list

**Supplementary Table 4**: Pairwise protein sequence alignment for patients with multiple major IGH clones

**Supplementary Table 5**: Pairwise protein sequence alignment for patients with multiple major IGL/K clones

**Supplementary Table 6**: WashU short predicted high quality neoantigen vaccine candidates

**Supplementary Table 7:** WashU predicted high quality fusion neoantigen vaccine candidates

**Supplementary Table 8**: WashU BCR long peptide vaccine candidates

**Supplementary Table 9**: BostonGene short predicted high quality neoantigen vaccine candidates

**Supplementary Table 10**: BostonGene predicted high quality fusion neoantigen vaccine candidates

**Supplementary Table 11**: WashU vaccine candidates selected for clinical trial

**Supplementary Table 12**: WashU vaccine candidates successfully manufactured

#### **Supplementary Table 13**: Clinical and metadata characteristics

**Supplementary Methods**

##### **Patient characteristics and sample acquisition:**

Four sets of patient samples were included. Sets 1-3 (53 patients) were cases retrospectively collected from the Washington University School of Medicine (WUSM) lymphoma banking program (sample IDs beginning with LYM). All patients provided written informed consent for the use of their samples in sequencing as part of the WUSM Lymphoma Banking Program. Excisional biopsy tissue and nonmalignant (skin punch biopsies) samples were collected (2008-2019). Pathology review was performed on frozen lymph node samples to confirm the diagnosis and estimate tumor cell content. Frozen sections (tumor and skin) were cut and used for genomic DNA isolation. Flow sorting was performed on a Reflection instrument. LYM520 RNA-Seq material was excluded due to RNA degradation; this was the only patient without RNA-Seq data. Patient set 4 (4 patients, 21 total samples) was prospectively collected for a pilot immunotherapy trial (Sample IDs beginning with FLNA). Tumors and uninvolved skin biopsies were obtained and flash frozen from consented (NCT03121677) individuals at Barnes-Jewish Hospital and reviewed to confirm the pathology diagnosis. All samples were collected within protocols approved by the WUSM institutional review board (201108251, 201804151) and all clinical characteristics are summarized in **Table S1**. PBMCs were obtained from four months post vaccination apheresis. The cells were isolated by standard Ficoll-paque Plus gradients and viably cryopreserved for later analysis.

##### **Library preparation and sequencing:**

Set 1: Genomic DNA was isolated using the QIAamp DNA Mini kit, with xylene. Library preparation, capture hybridization, and sequencing were performed as previously described.^27^ Briefly, DNA fragmentation was performed using the Covaris LE220 targeting an average fragment size of 200 bp. Dual indexed libraries were prepared using the KAPA HTP library prep kit (KAPA Biosystems) on the SciClone NGS instrument (Perkin Elmer). The final libraries were assessed using the LabChipGX (Perkin Elmer). Exome library capture was performed using the NimbleGen SeqCap EZ Exome v2.0 reagent. Sequencing was performed on a HiSeq 2000 with 2x100bp reads. These data completely overlap with our previously published study^1^.

Set 2: Genomic DNA was isolated using the QIAamp DNA mini kit. RNA was extracted using the Qiagen RNeasy extraction kit. DNA fragmentation was performed using Covaris LE220 targeting an average fragment size of 200 bp. Dual indexed libraries were prepared using TruSeq PCR-free (for WGS), and KAPA HTP (for WES), and TruSeq Stranded Total RNA (for RNA). DNA libraries were pooled and captured using the NimbleGen SeqCap EZ Exome v2.0 reagent or IDT exome kit. Sequencing was performed on a HiSeqX with 2x150bp reads for WGS, HiSeq 2500 with 2x125bp for RNA, and HiSeq 4000 with 2x150bp for WES. These data completely overlap with our previously published study^1^.

Set 3: For cases that were flow sorted, CD19+ and light chain restricted B cells were purified (>98% pure) using a Reflection instrument. Genomic DNA was isolated from purified cells using the QIAamp DNA mini kit. RNA was extracted using the Qiagen RNeasy extraction kit. DNA fragmentation was performed using Covaris LE220 targeting an average fragment size of 200 bp. Dual indexed libraries were constructed for DNA using the KAPA Hyper (Amplified) library prep kit on the SciClone NGS instrument. RNA libraries were constructed using Illumina TruSeq Stranded Total RNA Library Prep Gold. Corresponding (DNA and RNA) libraries were pooled and captured using an IDT exome kit. The final libraries were assessed using the LabChipGX. Sequencing was performed on Illumina HiSeq 4000 with 2x150bp reads. 15 normal samples were pooled/captured on 1 lane and 15 tumor samples were pooled/captured on 2 lanes for exomes. A total of 4 pools (5 libraries per pool) were captured on a total of 4 lanes (1 lane per pool) for RNA-Seq.

Set 4: Genomic DNA was isolated using the QIAamp DNA mini kit. RNA was extracted using the Qiagen RNeasy extraction kit. DNA and RNA were fragmented using the Covaris LE220 targeting an average fragment size of 200 bp. Dual indexed libraries were constructed for DNA using the Kapa Hyper (Amplified) library prep kit. RNA libraries were constructed using Illumina TruSeq Stranded Total RNA Library Prep Gold. The concentration of each library was determined through qPCR utilizing the KAPA library Quantification Kit. Corresponding (DNA and RNA) libraries were pooled and captured using an IDT Exome Capture Hybridization exome kit. DNA libraries were captured in one pool at a ratio of 60% tumor to 40% normal and sequenced on a NovaSeq6000 with 0.017 S4 flow cells with 2x150 bp reads. RNA libraries were captured separately and sequenced on a NovaSeq6000 with 0.010 S4 flow cell with 2x150 bp reads. Sample LYM193 is from previously published study^1^.

Sequencing data for Sets 1-4 have been deposited (dbGaP accession: phs001229).

##### **Parallel pipeline analysis:**

Two parallel analyses were conducted (referred to as WashU analysis pipeline and BostonGene analysis pipeline in the main text) with the same starting material of raw sequencing data (WGS, WES and RNA-Seq). The WashU analysis pipeline was used for the following analyses: 1) Mutational landscape depicted in **Figure 2, Table S2**; 2) Identification of fusion genes described in **Figure S1, Table S3**; 3) BCR clonality analysis depicted in **Figure S3a-f, Table S4-5**; 4) Neoantigen vaccine candidates discussed in **Figure 4,S4-6, Table S6-8**; 5) Clonality analysis **Figure S7-9**; 6) Clinical trial vaccine candidates described in **Table S10-11**. The BostonGene analysis pipeline was used for the following analyses: 1) BCR clonality analysis depicted in **Figure 3,S2,S3g-h**; 2) Neoantigen vaccine candidates discussed in **Figure S6, Table S9-10**; 3) Analysis of the MHC machinery described in **Figure 5**. Both pipelines were used to arrive at a final vaccine candidate list for the 4 patients enrolled in the pilot trial described in **Figure 6**.

#### **Retrospective neoantigen analysis**

##### Sequencing alignment and SNV/indel variant calling:

For sequencing data Sets 1-3 the WashU analysis pipeline first performed all alignments using the Genome Modeling System.^2^ Briefly, for exome data, paired-end reads were aligned to human reference sequence GRCh37, using BWA-MEM (v0.7.10)^3^, and de-duplicated using Picard (v1.8.5)^4^. Variants were identified using SAMtools (v0.1.18)^5^, SomaticSniper (v1.0.4), VarScan2 (v2.3.6), MuTect (v1.1.4), Strelka (v1.0.11), Pindel (v0.2.0), Breakdancer (v1.4.5) and GATK (v2.4.0), and annotated using the Variant Effect Predictor (VEP) (release 93.2) software for annotation of protein-coding effect of variants. Variants were filtered to remove common variants and pipeline artifacts. Manual review was performed as previously described.^6^ For RNA-seq data, FastQC (v0.10.0) and SAMStat (v1.0.8) were used to assess quality. Then the data was aligned to human reference sequence GRCh37 using Bowtie (v2.1.0) and TopHat (v2.0.8). Cufflinks (v2.1.1) was used to assemble transcripts, estimate their abundance and test for differential expression and regulation. BEDtools (v2.14.3) was then used to calculate read depth and create coverage plots.

For sequencing data Set 4 the WashU analysis pipeline first performed paired-end read alignment to human reference sequence GRCh38, using BWA (v0.7.15), and de-duplicated using Picard (v2.18.1) and SAMBLASTER (v0.1.24). Variants were identified using SAMtools (v1.3.1), VarScan2 (v2.4.2), Strelka (v2.9.9), Pindel (v0.2.5) and GATK (v3.6.0), and annotated using the Variant Effect Predictor (VEP) (release 93.2) software for annotation of protein-coding effect of variants. Variants were filtered to remove common variants and pipeline artifacts. Manual review was performed as previously described.^6^ For RNA-seq data, Picard, FastQC (v0.11.8) and Samtools (v1.3.1) were used to assess quality. Then the data was aligned to human reference sequence GRCh38 using HISAT2 (v2.0.5). StringTie (v1.3.3) and Kallisto (v0.43.1) were used to assemble transcripts (StringTie) and estimate their abundance.

The BostonGene analysis pipeline first aligned all exome data to the human reference genome GRCh38 (GRCh38.d1.vd1) using BWA v0.7.17. Duplicate reads were marked by Picard’s v2.6.0 MarkDuplicates, indels were realigned and base quality was recalibrated by GATK v3.8.1. Somatic single nucleotide variations (sSNVs), small insertions and deletions were identified using Strelka v2.9.

##### Identification of candidate fusion transcripts:

For sequencing data Set 1-3 the WashU analysis pipeline first aligned quality filtered reads using STAR-Fusion (v1.7.0) against transcriptome (GRCh37 Genecode v19 CTAT lib annotations) and genome (GRCh37) to identify fusion transcripts. Fusions were then filtered based on the following criteria: 1) Supported by at least 5 total reads (Junction plus Spanning reads), 2) Does not contain a pseudogene as a fusion gene partner, 3) Gene pairs were at least 1 Megabase apart. The remaining fusion candidates were then realigned using FusionInspector (v1.9.1) and alignment data was manually reviewed using Integrative Genomics Viewer (v2.7.0).

For sequencing data Set 4 the WashU analysis pipeline first aligned quality filtered reads using STAR-Fusion (v1.7.0) against transcriptome (GRCh38 Genecode v36 CTAT lib annotations) and genome (GRCh38) to identify fusion transcripts. Fusions were then filtered based on the following criteria: 1) Supported by at least 5 total reads (Junction plus Spanning reads), 2) Does not contain a pseudogene as a fusion gene partner, 3) Gene pairs were at least 1 Megabase apart. The remaining fusion candidates were then realigned using FusionInspector (v1.9.1) and alignment data was manually reviewed using Integrative Genomics Viewer (v2.7.0).

The BostonGene analysis pipeline detected gene fusions utilizing STAR-fusion v1.8.1. RNA-Seq data was aligned to the same genome by STAR v2.4.2 followed by realignment and base quality recalibration.

##### B-cell receptor alignment and analysis:

The WashU analysis pipeline consisted of the following steps: To determine a patient's B-cell receptor repertoire patient RNA-Seq data (fastq) was analysed with MiXCR (v3.0.2) with the following parameters: *analyze shotgun -s HomoSapiens --starting-material rna --receptor-type bcr --contig-assembly*. First, for all MiXCR clonotype results (files ending with “.clonotypes.IGH.txt“), including the aaSeqFR3, aaSeqCDR3 and aaSeqFR4 amino acid sequences were concatenated. Next, the counts for these concatenated amino acid sequences were collapsed if identical or entirely contained within a longer sequence, and total clone count summed. All immunoglobulin heavy (IgH) chain and immunoglobulin light/kappa (IgL/K) chain clonotypes were then separated into two groups and their corresponding clone count was then used to recalculate their respective clone fraction. Clonotypes with clone fractions greater than or equal to 9% were labeled as clonal B-cells.

The BostonGene analysis pipeline consisted of the following steps: BCR profiling was conducted using MiXCR v3.0.12 from RNA-Seq reads using default options for shotgun sequencing. To capture somatic hypermutations in malignant B-cells all the related BCR clonotypes sharing the same V(D)J segments and CDR3 regions differing by no more that two nucleotides were combined into clonotype groups. Normal BCR structures were obtained from publicly available bulk RNA-Seq samples with high B-cell population according to the tissue of origin, in total 110 samples. BCR profiling was performed as described above. Only high covered samples with more than 100 assembled clonotypes in each of the chains (heavy, kappa and lambda) were kept for further analysis. Publicly available datasets used: GTEx [phs000424](https://www.ncbi.nlm.nih.gov/projects/gap/cgi-bin/study.cgi?study_id=phs000424) (N=15)^7^, GSE90081 (N=12)^8^, GSE112057 (N=12)^9^, GSE61410 (N=4)^10^, GSE111405 (N=3)^11^, GSE63816 (N=3)^12^, GSE58335 (N=2)^13^, GSE43603 (N=1)^14^, GSE57944 (N=1)^15^.

##### Vaccine candidate prioritization and analysis:

The WashU analysis pipeline consisted of the following steps: To determine a patient’s HLA class I allele type we used OptiType (v1.3.3) on normal WES data. Epitope predictions were performed for each patient’s mutation(s) against all 6 of their predicted HLA class I alleles using pVACtools (v1.5.3)^16^. All patient-predicted peptides were condensed to the single best high quality candidate based on binding affinity. Small somatic variant neoantigen candidates (pVACtools pVACseq results) were further filtered using the following criteria: (1) HLA binding affinity IC50 ≤ 500 nmol/L (Note: peptides of sizes 8-11, with a sliding window around the mutation, using a number of binding prediction algorithms, and then calculated a median binding affinity); (2) normal/tumor DNA/RNA coverage > 10; (3) normal DNA variant allele frequency (VAF) < 1%; (4) tumor DNA/RNA VAF > 5%; and (5) gene expression FPKM > 1. B-cell clonotype neoantigen candidates (pVACtools pVACbind results) were filtered to identify high quality candidates (HQC) using the following criteria: (1) B-cell clonotype percentage ≥ 9% and (2) HLA binding affinity IC50 ≤ 1,000 nmol/L. Gene fusion neoantigen candidates (pVACtools pVACfuse results) were filtered to identify HQC using the following criteria: (1) A minimum of 5 total junction and/or spanning reads and (2) HLA binding affinity IC50 ≤ 500 nmol/L.

The BostonGene Vaccine Module V1 reconstructs candidate tumor specific peptides by assembling mutant transcripts from tumor RNA reads. Mutant transcripts are identified as those supporting somatic variants previously evaluated from exome data. The Module uses IsoVar v0.7.0 to reconstruct such peptides. HLA genotypes were identified from RNA-Seq data using seq2hla v2.2. Peptide-HLA affinity is evaluated by a custom prediction model. Briefly an allele-specific gradient boosting model was trained on curated data from IEDB and validated on independent datasets. Peptide encoding scheme included 1-hot encoding and peptide features such as mass, isoelectric point, hydrophobicity, instability index according to Guruprasad et al, 1990 and flexibility according to Vihinen, 1994. Fusion derived neoantigens were predicted by INTEGRATE-Neo (v1.2.1) from the events identified by STAR-fusion (v1.8.1) using NetMHC 4.0 for neoantigen-HLA affinity evaluations. Fusions should have been supported by at least 10 junction reads and neoantigens-HLA IC50 should be under 500nM.

##### Evaluating efficiency of antigen presenting machinery:

To evaluate antigen presenting machinery (APM) status in FL cells we assessed for somatic aberrations in genes involved in APM function and analyzed expression levels of HLA class I & II genes. We identified non-synonymous mutations and gene fusions in all samples (**Figure 5a**) for 32 manually selected genes known to be involved in APM. Gene fusions were calculated by STAR-fusion, small variants - by Strelka (see Sequencing alignment and variant calling above for details). To correctly evaluate polymorphic HLA gene expression we replaced HLA transcripts in GENCODE 33 assembly with an IMGT set of HLA transcripts and used kallisto to quantify transcript abundance for the samples. Then we normalized expression by log-transform followed by median-transform scaling within each of the batches (**Figure 5b**).

##### Evaluating clonal and subclonal architecture of FL samples

We used SciClone (v1.0.7)^17^ for single-region subclonal reconstruction. Input VCFs were used to calculate variant allele frequencies and CNA inputs were used to determine regions with loss of heterozygosity. Only SNVs in clonally copy number neutral (major = 1, minor = 1) regions with no subclonal CNAs were considered by SciClone (v1.0.7) and all samples were run using default parameters. Mutation clusters defined by SciClone (v1.0.7) were characterized using variant allele frequencies.

#### **Pilot trial neoantigen analysis**

##### Sequencing alignment and SNV/Indel variant calling:

The same analysis strategy was used as described above for the retrospective neoantigen analysis.

##### B-cell receptor alignment and analysis:

The same analysis strategy was used as described above for the retrospective neoantigen analysis, with one exception: Clonotypes with clone fractions greater than or equal to 9% were labeled as clonal B-cells; however, if no clonal B-cell populations could be identified than the largest IgH and IgL/K clonotypes were labeled as the patient’s clonal B-cells.

##### Vaccine candidate prioritization and analysis:

The WashU analysis pipeline consisted of the following steps: To determine a patient’s HLA class I allele typing we used OptiType (v1.3.3)^18^ on normal WES data. Epitope predictions were performed for each patient’s mutation(s) against all 6 of their predicted HLA class I alleles using pVACtools (v1.5.3). All patient-predicted peptides were condensed to the single best high quality candidate (HQC) based on binding affinity. Small somatic variant neoantigen candidates (pVACtools pVACseq results) were further filtered using the following criteria: (1) HLA binding affinity IC50 ≤ 500 nmol/L; (2) normal/tumor DNA/RNA coverage > 10; (3) normal DNA variant allele frequency (VAF) < 10%; (4) tumor DNA/RNA VAF > 5%; and (5) gene/transcript expression FPKM ≥ 1. However, to achieve the maximum number of vaccine candidates (20, as stipulated by manufacturer) possible per patient, the previous filtering criteria were relaxed in reverse order creating 6 tiers. Tier1 candidates passed all filtering criteria. Tier2 candidates ignored filtering criteria 5 only. Tier3 candidates ignored filtering criteria 4 only. Tier4 candidates ignored filtering criteria 3 only. Tier5 candidates ignored filtering criteria 2 only. Tier6 candidates ignored filtering criteria 1 only. The same BostonGene Vaccine Module V1 strategy used within retrospective neoantigen analysis was also applied to trial patient data. The union of both methods (WashU and BostonGene analysis pipelines) were used together to ensure that no potentially important candidates were missed due to differences of analysis. If a vaccine candidate was identified by the BostonGene analysis pipeline and not by the WashU analysis pipeline, then an investigation was conducted to evaluate the missing candidate and determine if it should be included within the patient’s vaccine. The largest IgH and IgL/K B-cell clonotype population’s CDR3 regions were considered high quality neoantigen vaccine candidates.

##### Clinical trial design and approval:

To assess the feasibility and safety of a personalized neoepitope SLP (synthetic long peptide) vaccine in combination with nivolumab, we initiated a pilot trial (IRB and FDA IND-approved, open to accrual, NCT03121677, **Figure S10**). Vaccine was administered in combination with polyIC:LC (Hiltonol), a TLR3 agonist immune adjuvant, and nivolumab (anti-PD-1 monoclonal antibody), in four limb specific pools (described above) at the study-designated time points. Patients who progress on post-cycle 2 or post-cycle 6 restaging were allowed to receive rituximab in combination with vaccine and nivolumab in an effort to keep patients safely on trial and still meet the critical correlative time points. As rituximab does not affect T cell responses, and we are not expecting humoral responses from the neoantigen vaccines, we do not expect addition of rituximab to interfere with the efficacy or interpretation of vaccine response. A detailed chronological treatment schema is shown in **Figure S11**. Eligible patients must have had relapsed FL which had been previously treated with an anti-CD20 antibody and an alkylator. Patients were ineligible if they were refractory to rituximab (as defined as no response to or progression of FL <6 months following prior anti-CD20 mAb therapy), required systemic anti-lymphoma therapy within the last six months, had a history of transformed follicular lymphoma, or had prior therapy with PD-1 or PD-L1 inhibitors. Patients were required to have a tumor accessible for biopsy and were deemed to be appropriate for initiation of the next line of therapy for 4 to 5 months. Primary outcome measures of the trial are feasibility and safety of the neoantigen vaccine in combination with nivolumab as measured by the number of participants whose personal vaccines can be manufactured and delivered without unacceptable toxicity. Unacceptable toxicity is defined as the inability to receive further therapy due to toxicities of therapy as defined by NCI Common Terminology Criteria for Adverse Events (CTCAE) version 4.03 or the occurrence of other toxicities deemed to be at sufficiently high risk to patients by the principal investigator. Secondary outcome measures include overall response rate (ORR), complete response (CR) rate, duration of response, progression-free survival (PFS), overall survival (OS), and partial response (PR) rate.

##### Vaccine candidate design, pooling strategy and manufacturing

All short and BCR peptide vaccine candidates were BLASTp ^19^aligned against the human reference genome to ensure that the neoantigen vaccine candidate did not match any other portion of the human genome, to avoid off target-effects. Next, all somatic short peptides (8-11 amino acids) were required to have a long peptide (25 amino acids) register, with preservation of the short peptide after proteasome processing by NetChop (v. 3.1)^20^. Additionally, inclusion of multiple long peptide registers were selected for a given somatic short peptide when possible. All BCR long peptide (25 amino acids) registers were selected by subtracting the BCR CDR3 vaccine candidate amino acid lengths from the total and then including the difference of amino acids given the flanking FR3 and FR4 corresponding to a left (inclusion of FR3 flanking amino acids only), middle (equal inclusion of FR3 and FR4 amino acids) and right (inclusion of FR4 flanking amino acids only). Long peptide N- and C-terminus sequence recommendations (generally, do not start or end with a bulky amino acid): avoid Glutamine (Q) at the N-terminals and avoid Cysteine (C) and Proline (P) at the C-terminals. Additionally, all long peptide vaccine candidates were BLASTp aligned against the human reference genome to ensure that the neoantigen vaccine candidate did not match any other portion of the human genome to avoid off target-effects. GMP grade vaccinating peptides were obtained via Neon Therapeutics (Cambridge, MA) and synthesized by CreoSalus (Louisville, KY) using standard solid-phase synthetic chemistry and then purified using reverse phase-HPLC. Up to twenty long peptides were combined into four pools, minimizing competition for MHC, taking into consideration predicted binding affinities, for limb specific injection. Immediately before inoculation, pools were emulsified with Hiltonol (Poly-ICLC), Oncovir, Inc by the Siteman Cancer Center Clinical Pharmacy.

##### Identification of candidate encoding peptides binding to HLA A*68:01 for patient FLNA-04

A TAP deficient cell line was created by expression of ICP-47 (gift from Ted Hansen) under puromycin selection in the MHC class I negative, lymphoblastoid cell line, 721.221 (gift fromM. Colonna, Washington University Saint Louis). A TAP negative clone was transduced with synthetically constructed, HLA A*68:01 in a IRES GFP retrovirus. The greater than 95% GFP positive cell line was serum starved for 1 hour prior to plating with various concentrations of in silico predicted candidate short peptides synthesized by GenScript to greater than 90% purity. Peptides corresponding to candidate nonomers (9-mers) were solubilized in 10% DMSO and then incubated with the Class I specific/TAP deficient cell line for three hours at ambient temperature in RPMI1640 containing 100 mM HEPES, 2mM glutamine, 1000 un/mL Penicillin, 1mg/mL Streptomycin, and 1% (v/v) heat inactivated human AB serum. The cells were shifted to 37C for overnight incubation then washed twice and stained with fluorescently labeled W6/32 monoclonal antibody to detect Class I expression. Cells were washed twice and then collected on a Beckman Coulter Navios flow cytometer utilizing 7-AAD viability dye to exclude dead cells from later cell analysis. Flow cytometric data was analyzed on FlowJo v. 10.6.1. Mean fluorescence intensity (MFI) of 7-AAD negative/GFP positive/W6/32 positive cells was determined in duplicate for each peptide concentration. **Figure 6C**, shows the MFI over baseline (cells with no peptide) for the candidates that stabilized peptides relative to the FluNP 91-99 positive control and CTIF D459G negative control peptide. The red line shows baseline W6/32 MFI of the highest concentration of CTIF D459G negative control peptide. The results shown are the average of three independent determinations.

##### ELISPOT

Two million PBMCs from four month post vaccination were stimulated with vaccine synthetic long peptides (SLP) at 40 ug/mL in complete OpTmizer T-cell media (Gibco) supplemented with 5% human AB serum. Cells were fed every other day with 50 um/mL IL-2 (Proleukin) until day ten and then used in a standard IFN- ELISPOT assay (Cellular Technologies Limited, OH) using the predicted minimal short peptide candidates for re-stimulation. Developed plates were counted and analyzed blindly by the Immunomonitoring lab at the Bursky Center for Human Immunology and Immunotherapy Center, Washington University) on a C.T.L.-Immunospot S6 Universal Analyzer. Results were de-convoluted and shown in **Figure 6E** as spots per 1e6 PBMC.

##### Tetramer Assay

Day ten stimulated PBMCs used in the ELISPOT were stained with freshly prepared HLA A*68:01 tetramers containing the minimal short peptide (SP) candidates fluorescently labeled with PE and APC for 30 min at ambient temperature. Additionally, 7-AAD-/CD19-/CD56-/CD3+/CD8+ T cells double positive for tetramer PE and tetramer APC were considered antigen specific T cells.

##### Intracellular Cytokine Assay

Class I negative K562 leukemic cells were transduced with HLA A*68:01 with GFP and sorted to >95% purity and used as an artificial antigen presenting cell (AAPC). AAPC were loaded with short peptide of interest for one hour, washed, irradiated and then mixed at a 5:1 (E:T) ratio with day ten SLP stimulated PBMCs. Cells were treated with BrefeldinA and monensin and intracellular cytokine was allowed to accumulate for six hours. Cells were then stained for IFN- and TNF as previously described.^21^ After six hours, cells were stained with Zombie green (ZG) (Biolegend) to remove dead cells from the analysis and CD45+/CD3+/CD8+ cells were examined for expression of the inflammatory cytokines IFN-g and TNF. Data was collected on a BeckmanCoulter Navios Flow Cytometer and analyzed on FlowJo (v10.6.1).

### **Supplementary Figures**

#### **Supplementary Figure 1: Manually reviewed fusions observed for all patients**

The bar graph on the top corresponds to the number of total fusions per patient and colored by fusion type. The bar graph on the left corresponds to the percentage of fusions for a given gene for the entire cohort. Only a single fusion transcript pair is recognized per gene pair even if a patient has multiple fusion transcript pairs within that fusion.

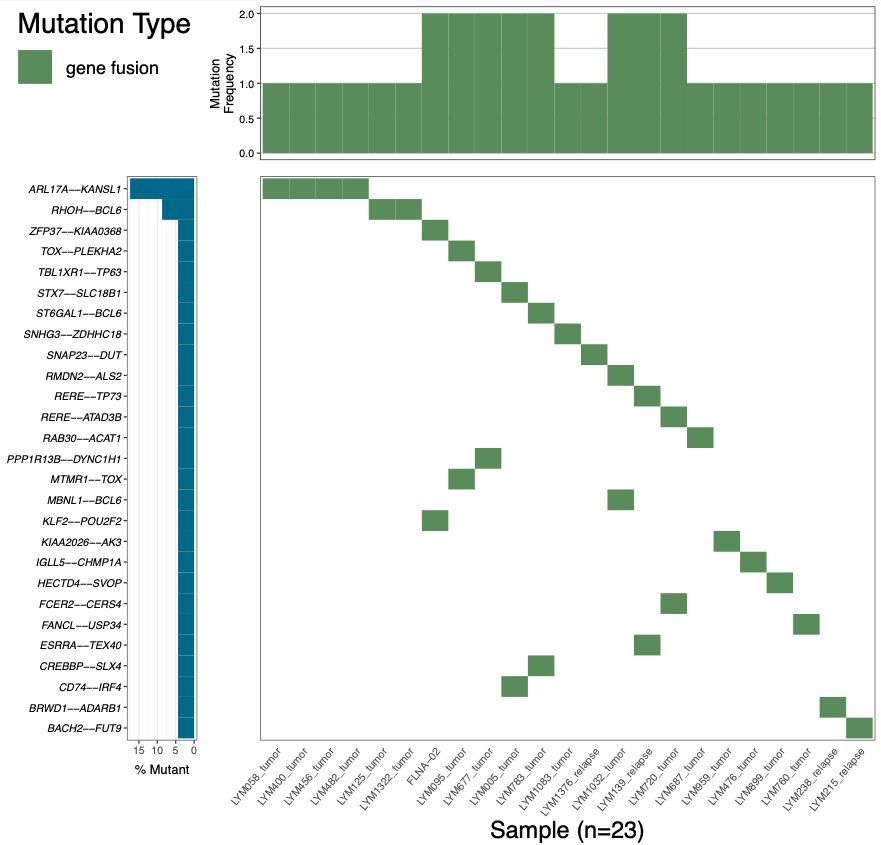

#### **Supplementary Figure 2: Comparison of healthy and malignant B-cell repertoires**

(a) Overall RNA-Seq coverage met minimum quality metrics (e.g., >50M reads) for all the cohorts of both healthy/normal and tumor (FL) samples. (b) BCR clonality evaluated individually by heavy, kappa and lambda chain repertoires that are much higher in tumor than normal samples. (c) Prevalent clonotype in tumor samples also had much higher content compared to normal samples being evaluated for all three chains. *** denotes p-value = 0.001.

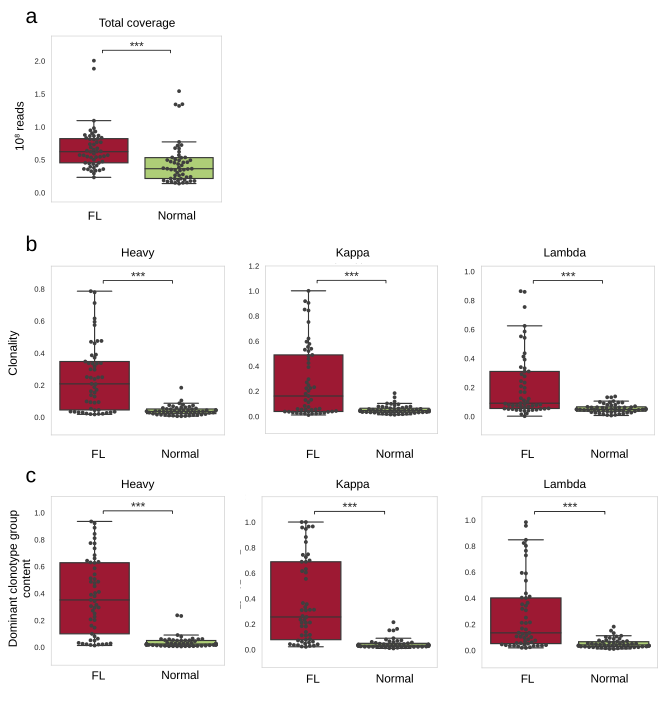

#### **Supplementary Figure 3: B-cell tumor repertoire oligoclonality analysis identifies oligoclonal and polyclonal populations within follicular lymphoma patients**

Heatmaps of B-cell Receptor repertoire clonality for (a) immunoglobulin heavy chain (IgH) and (b) for immunoglobulin light/kappa chain (IgL/K) were generated by MiXCR and include only FR3, CDR3, and FR4 regions. Clonotype clonal fractions are displayed as colours ranging from white to purple as shown in the key. (c) The total number of reads used in clonotype generation. Heatmaps of best median IC50 score for major clonotypes for (d) IgH and (e) for IgL/K scores were generated by the pVACtools pVACbind module using trimmed regions of FR3 and FR4 which span CDR3. The median IC50 scores are displayed as colours ranging from red to blue as shown in the key. Protein sequences with a gap or stop codon were excluded entirely. (f) The total number of BCR vaccine candidates predicted per patient. (g) The table contains the 9 most abundant B-cell clonotypes for patient sample LYM120_t and the highlighted nucleotide colors indicate whether a nucleotide matches the majority of the alignment (orange) and if a mis-match exists (red). (h) Each circle graph represents a patient’s entire B-cell repertoire and if B-cells are subclones determined via alignment they are linked through blue lines. For a, b, d, and e, Only the first 5 major clonotypes are displayed for all patients and the entire cohort is sorted by IgH clonality.

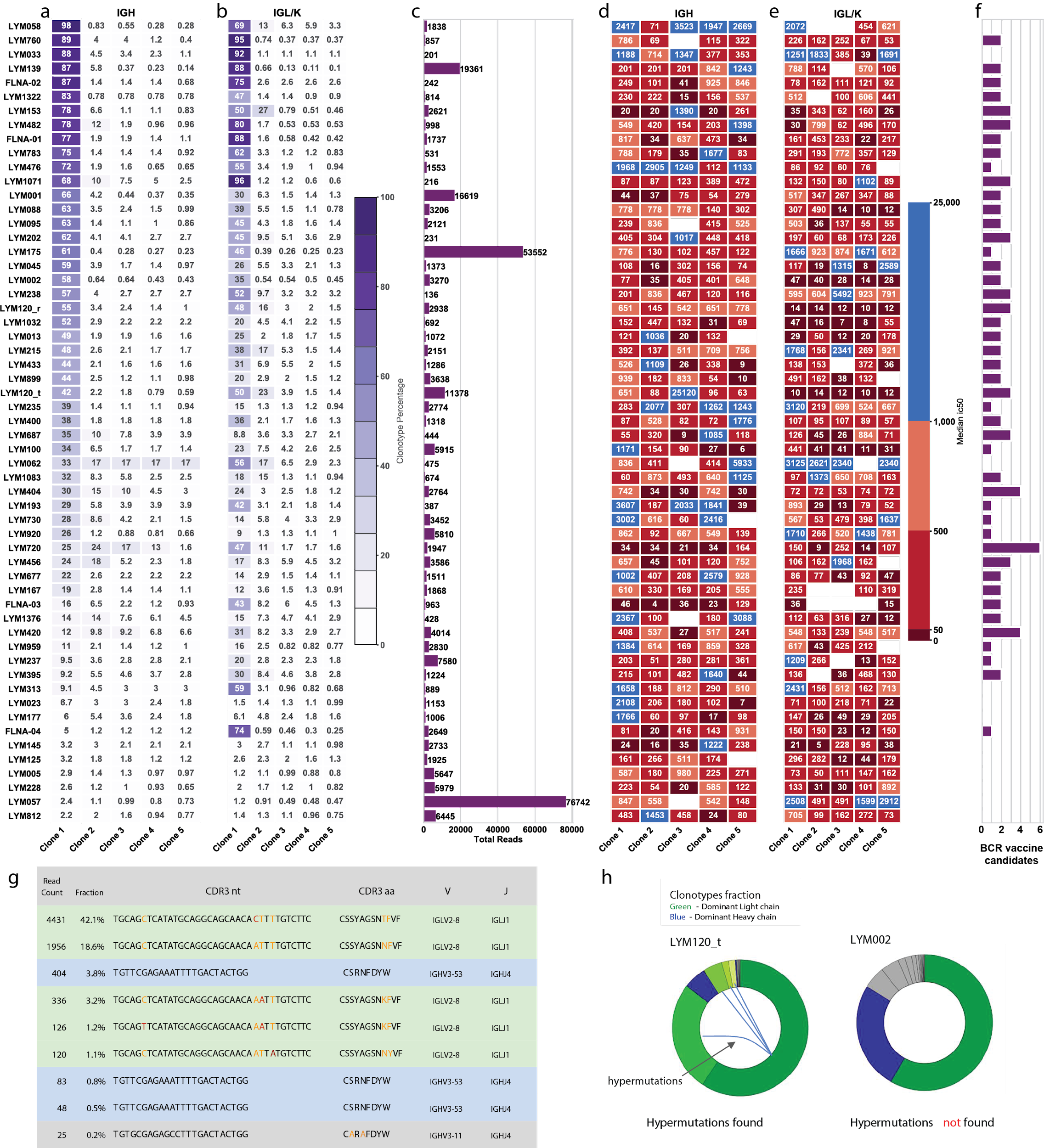

#### **Supplementary Figure 4: Recurrently mutated genes and mutation burden observed for all predicted high quality neoantigens**

The bar graph on the top corresponds to the number of total mutations per patient predicted as high quality neoantigens and colored by mutation type. The bar graph on the left corresponds to the percentage of mutations for a given gene for the entire cohort. Columns represent each patient in the cohort and are ordered by the presence of mutations in the most to least frequently mutated gene. Only a single mutation is recognized per gene even if a patient has multiple mutations within that gene, priority order is indicated in the legend from top to bottom.

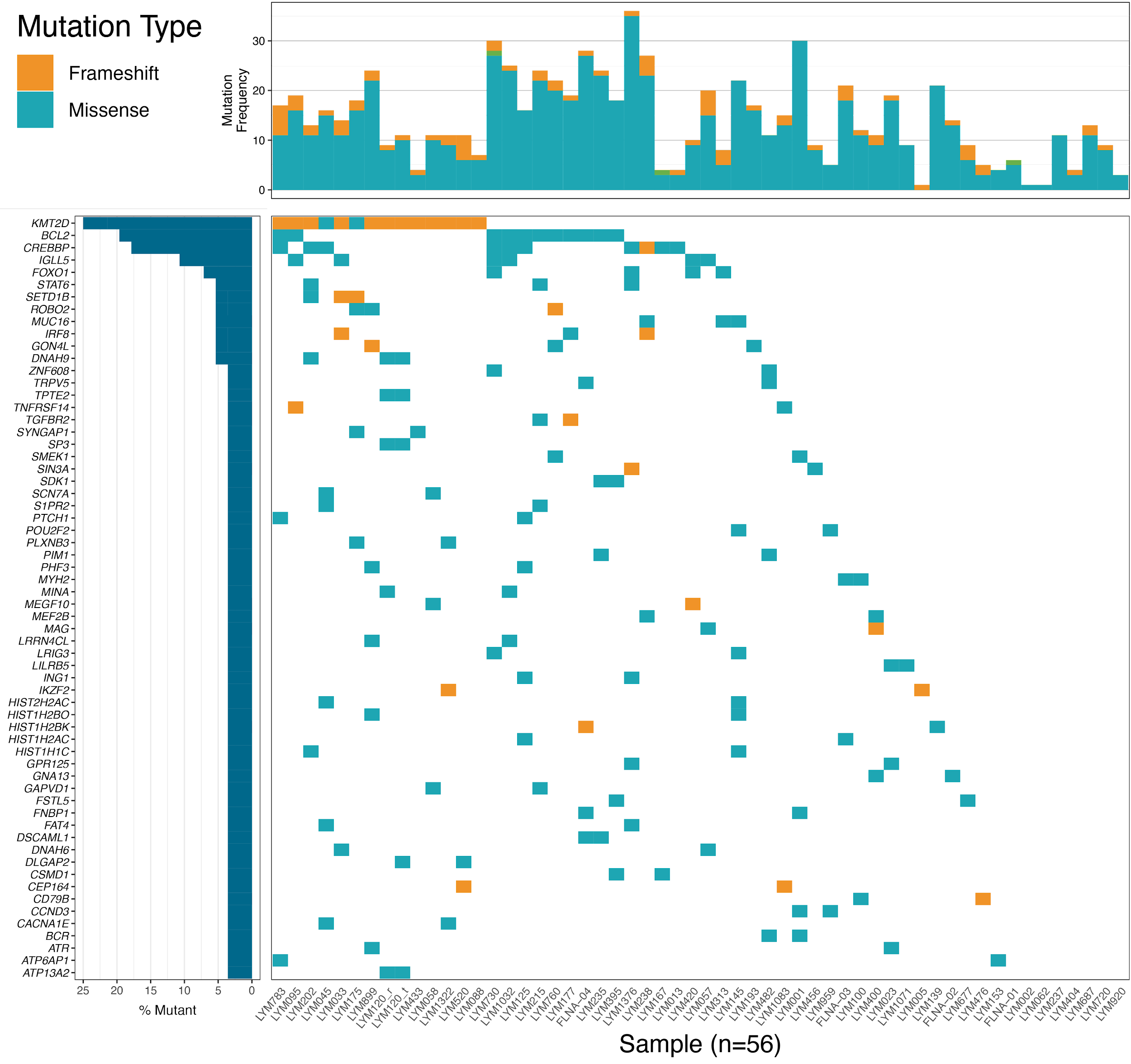

#### **Supplementary Figure 5: Fusions predicted to result in high quality neoantigen vaccine candidates**

The bar graph on the top corresponds to the number of total fusions per patient and colored by fusion type. The bar graph on the left corresponds to the percentage of fusions for a given gene for the entire cohort. Only a single fusion transcript pair is recognized per gene pair even if a patient has multiple fusion transcript pairs within that fusion, priority order is indicated in the legend from top to bottom.

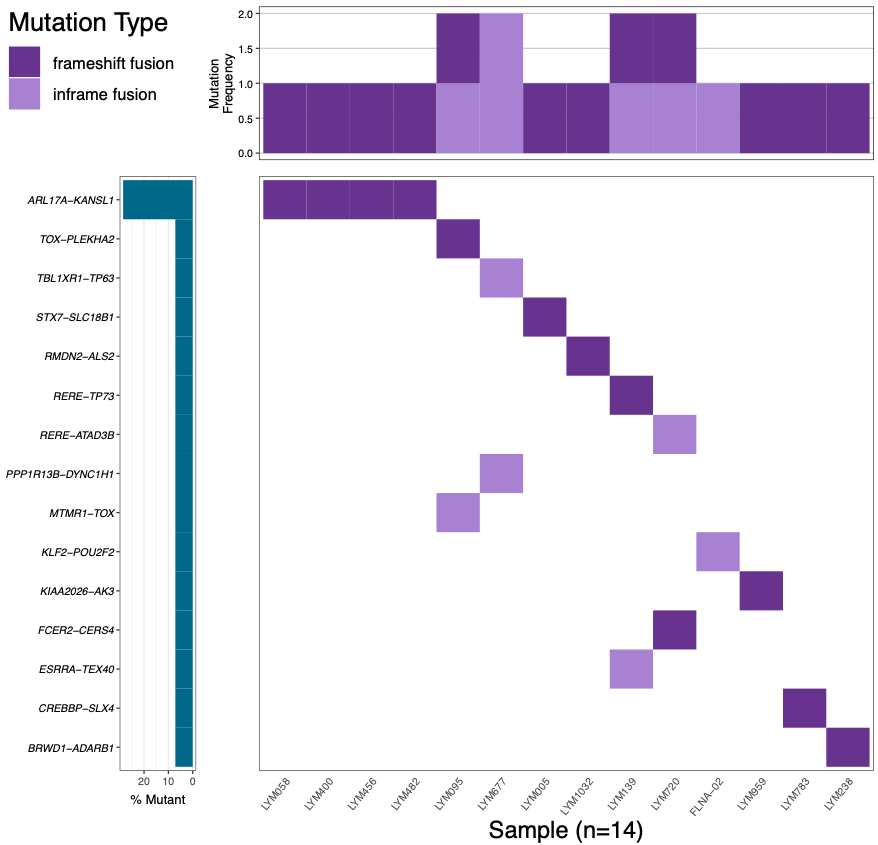

#### **Supplementary Figure 6: Predicted high quality neoantigen predictions and comparison between WashU and BostonGene analysis pipelines**

Concordance of complementary approaches for neoantigen prediction. The bar graph shows the number of neoantigens unique to and shared between the WashU analysis pipeline and the BostonGene Vaccine Module V1 pipeline.

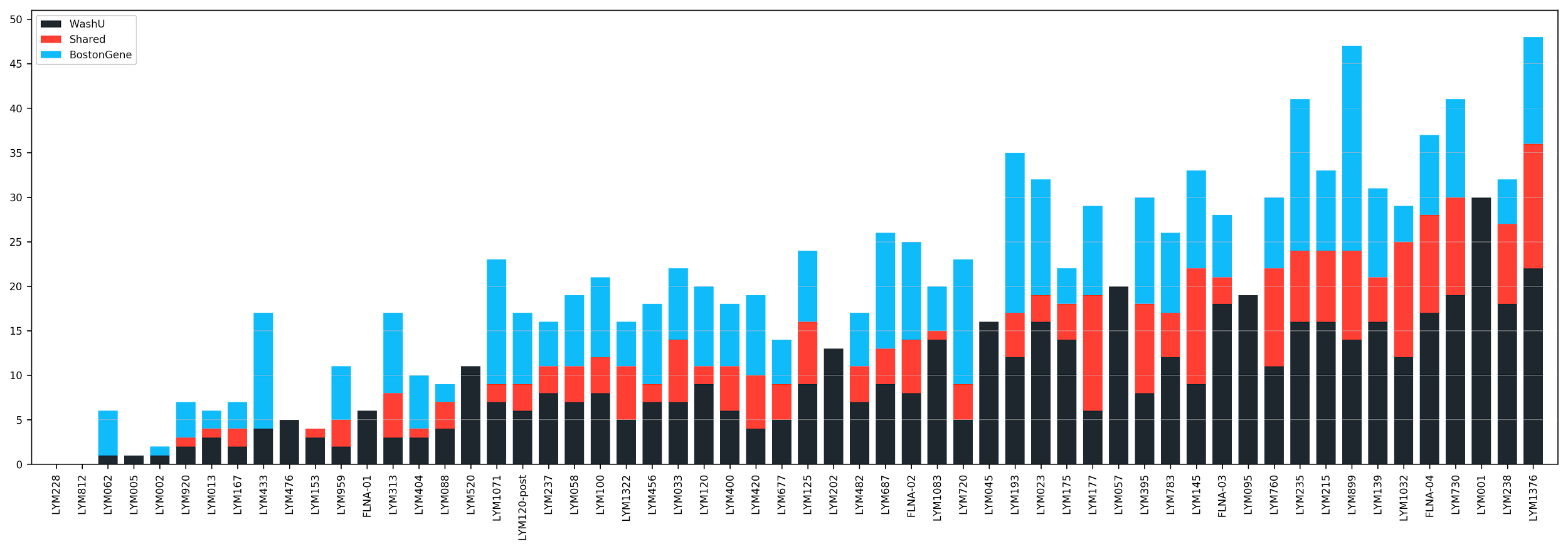

#### **Supplementary Figure 7: Distribution of read depths for all manually reviewed variants**

Boxplots of each individual patient read depths for all manually reviewed variants within that patient.

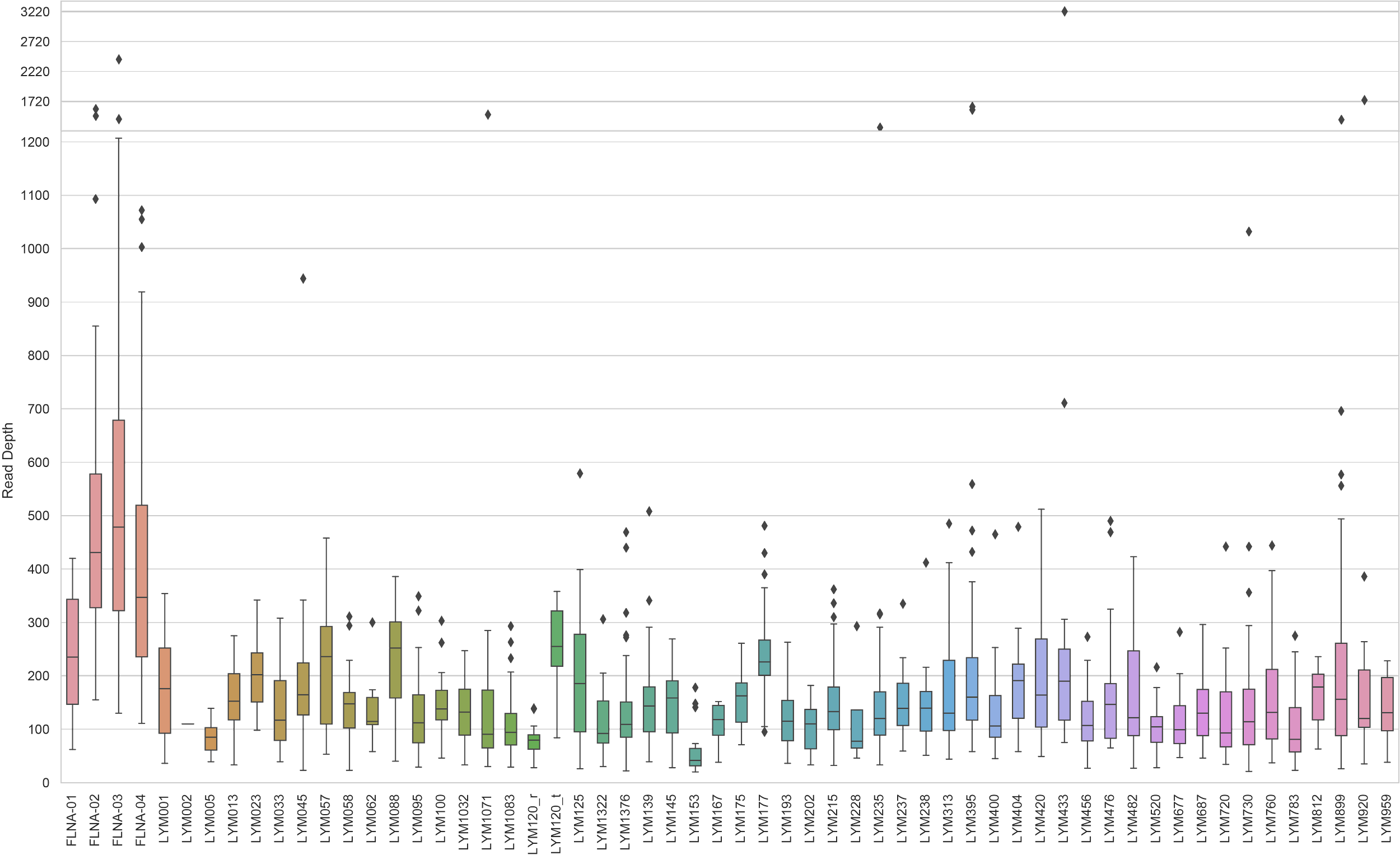

#### **Supplementary Figure 8: Distribution of DNA VAFs for all manually reviewed variants**

Boxplots of each individual patient tumor DNA VAF for all manually reviewed variants within that patient.

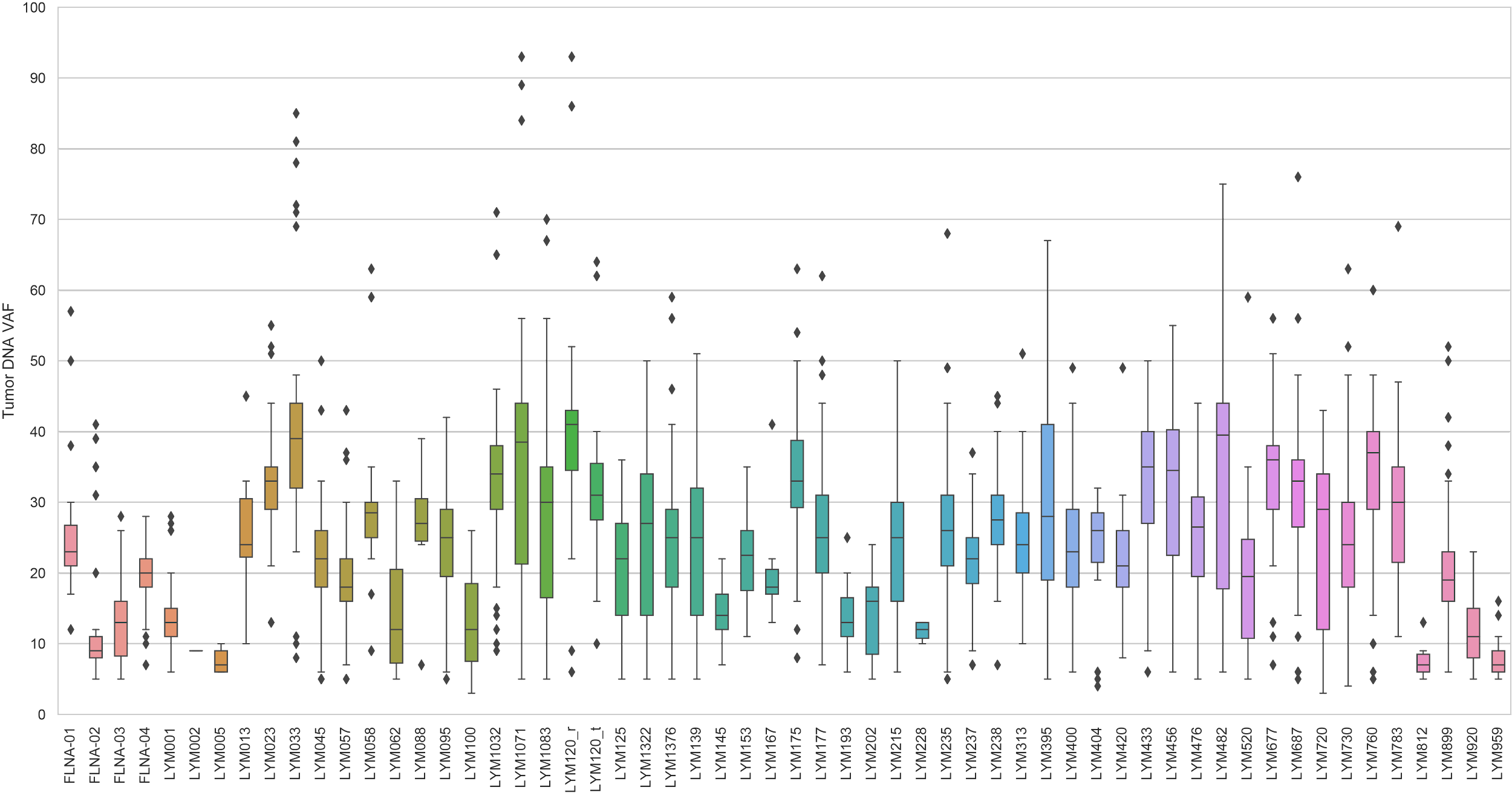

#### **Supplementary Figure 9: Inferred subclonal architecture for all patients**

Each patient’s SciClone plot is made up of two graphs: 1) Kernel density plots of VAFs across regions with copy number two, posterior predictive densities summed over all clusters for copy number neutral variants, and posterior predictive densities for each cluster/component and 2) Clusters detected using variants from copy number neutral segments.

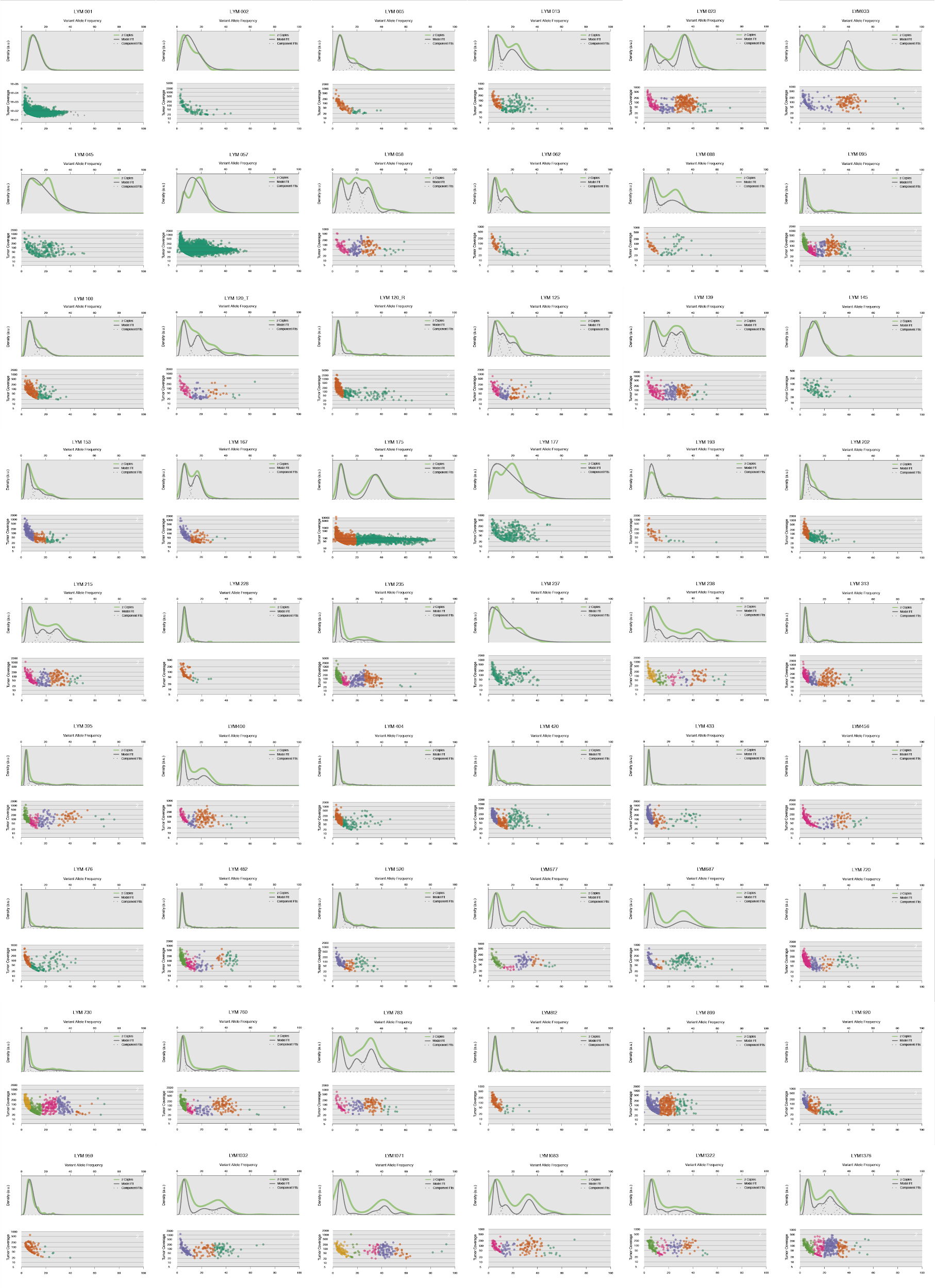

#### **Supplementary Figure 10: Clinical trial treatment schema (#NCT03121677)**

Treatment schema showing Day, Week, and Cycle of all staging, treatment biopsy, blood, draw, and leukapheresis events. Note that rituximab is only administered if the patient experiences progressive disease while awaiting vaccine study therapy.

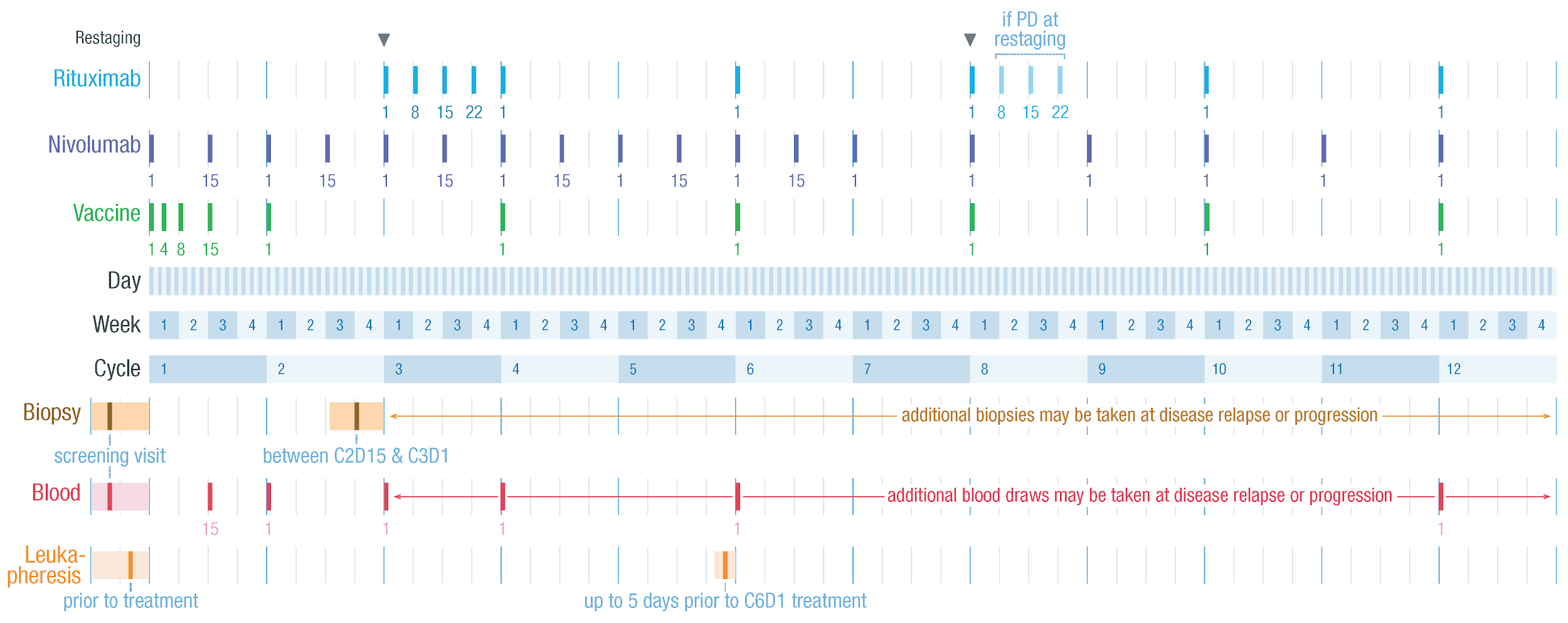

#### **Supplementary Figure 11: Overview of pilot trial to evaluate neoantigen vaccine + ICT in FL**

Patients with relapsed FL undergo a study related biopsy which is used for synthetic long peptide vaccine design following exome sequencing (tumor/normal), transcriptome sequencing (tumor) and neoepitope prediction. If neoepitopes are identified a vaccine is manufactured and administered in combination with polyIC:LC (Hiltonol) and nivolumab. Patients who progress on post-cycle 2 or post-cycle 6 restaging are allowed to receive rituximab in combination with vaccine and nivolumab in an effort to keep patients safely on trial and still meet the critical correlative time points. Primary outcome measures of the trial are feasibility and safety of the neoantigen vaccine in combination with nivolumab as measured by the number of participants whose personal vaccines can be manufactured and delivered without unacceptable toxicity. Secondary outcome measures include overall response rate (ORR), complete response (CR) rate, duration of response, progression-free survival (PFS), overall survival (OS), and partial response (PR) rate. Immune response to predicted neoantigens was assessed for MHC-allele-specific binding and T cells activation by ELISPOT, MHC tetramer-based assays, and high dimensional profiling (CyTOF) of CD4+ and CD8+ T cells at time points before, during and after therapy.

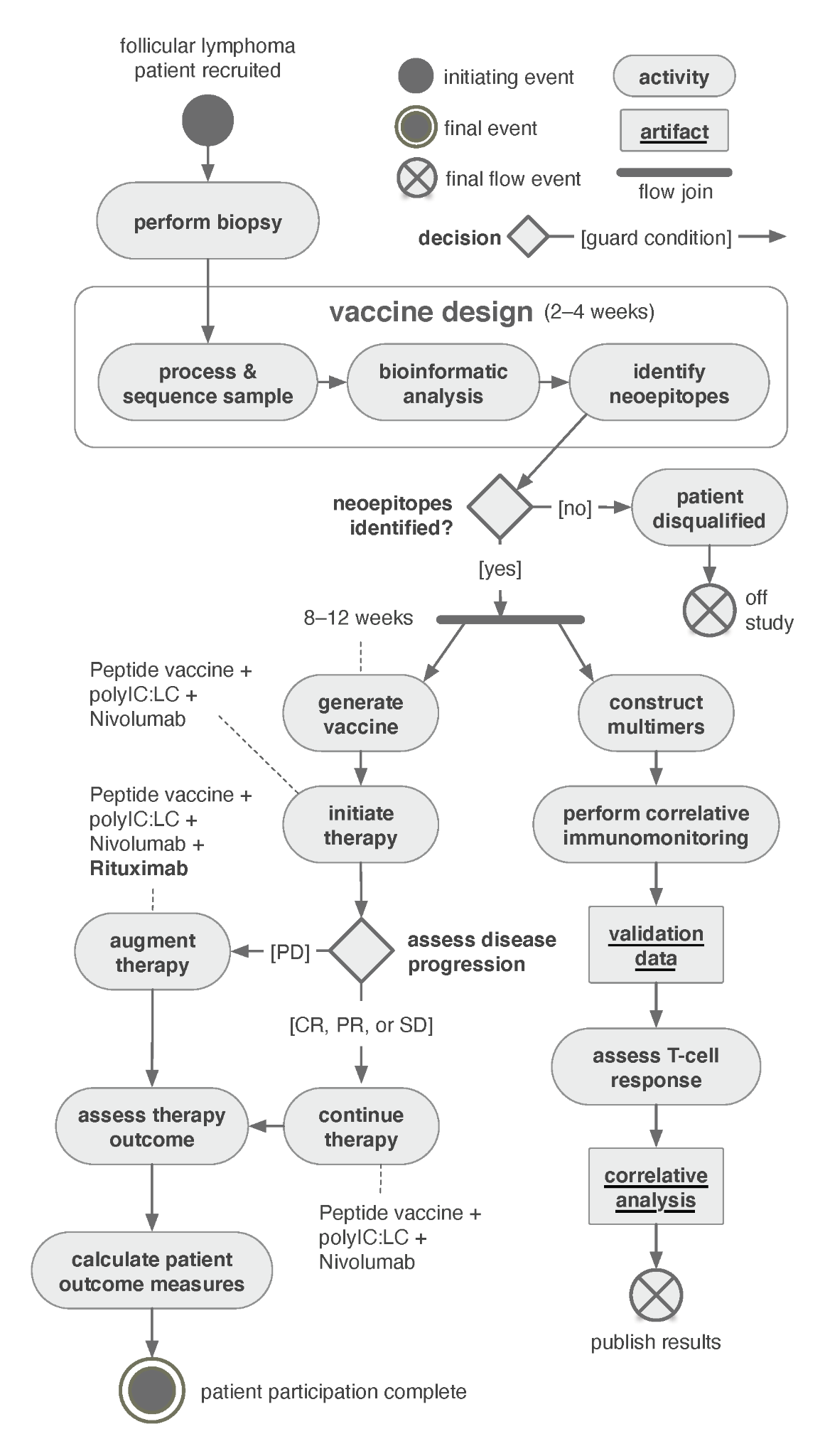

### **Supplementary Tables**

#### **Supplementary Table 1: Clinical characteristics of patients used for genetic analysis**

| **Characteristics** | **Genetic analysis values** |
| --- | --- |
| Total number of patients | 57 |
| Female, % | 46 |
| Male, % | 54 |
| Age (Median) | 55 |
| Age range | 22-77 |
| **Stage, %** |  |
| I | 12 |
| II | 5 |
| III | 37 |
| IV | 46 |
| **FLIPI score, %** |  |
| Low | 37 |
| Intermediate | 26 |
| High | 33 |
| NA: no information | 4 |
| **Lymphoma type, %** |  |
| FL | 84 |
| Transformed lymphoma (tNHL) | 16 |
| **Sequenced biopsy, %** |  |
| Treatment-naïve FL | 47 |
| Relapsed FL | 37 |
| Transformed lymphoma (tNHL) | 16 |

#### **Supplementary Table 2: VCF file containing WashU manually reviewed short somatic variants**

See [Supp](https://drive.google.com/file/d/1sMqCYgNRamG4c1oqRUvtC45mIOJIqLNc/view?usp=sharing) Table2.tsv

#### **Supplementary Table 3: WashU manually reviewed gene fusion list**

See Supp Table3.xlsx

#### **Supplementary Table 4: Pairwise protein sequence alignment for patients with multiple major IGH clones**

See Supp Table4.png

#### **Supplementary Table 5: Pairwise protein sequence alignment for patients with multiple major IGL/K clones**

See Supp Table5.png

#### **Supplementary Table 6: WashU predicted high quality neoantigen vaccine candidates from small variants**

See Supp Table6.tsv

#### **Supplementary Table 7: WashU predicted high quality neoantigen vaccine candidates from fusions**

See Supp Table7.xlsx

#### **Supplementary Table 8: WashU long peptide vaccine candidates from major BCR clone(s)**

Long peptide sequences which could be used to potentially treat the patients. At least one IGH and IGL/K major clone was selected (which has a minimum clonality of 9% and has a minimum clone count of 5, otherwise the major clone is selected by default) for every patient based on the best median ic50 score reported by pVACbind results.Thereotherically, these long peptide sequences would be manufactured and used as a portion of the personalized cancer vaccine for the individual patients.

See Supp Table8.xlsx

#### **Supplementary Table 9: BostonGene short predicted high quality neoantigen vaccine candidates**

See Supp Table9.tsv

#### **Supplementary Table 10: BostonGene predicted high quality fusion neoantigen vaccine candidates**

| **Patient** | **Fusion** | **Epitope** | **Affinity, nM** | **HLA allele** |
| --- | --- | --- | --- | --- |
| LYM013 | ARHGEF18--CD320 | VLYGTNEIL | 208.02 | A02:01 |
| LYM139 | PTPRC--NBPF14 | FLDTEVFVTV | 5.97 | A02:01 |
| LYM1032 | RMDN2--ALS2 | GGFPQALKK | 158.48 | A11:01 |
| FLNA-02 | KLF2--POU2F2 | HLRTHTEIR | 56.46 | A31:01 |
| LYM005 | MTG1--SMAD7 | KQIPNFFWSL | 75.88 | A02:01 |
| LYM013 | CNN2--PPP1R12C | ALAGKLRNQK | 49.8 | A03:01 |
| LYM023 | ELK3--CDK17 | YYYDKSLLX | 19.49 | A29:02 |
| LYM045 | TPM4--CTD-2192J16.20 | KAADESERTQW | 64.79 | B57:01 |
| LYM058 | CAPZA2--DYRK1A | EEKVIVLPL | 164.23 | B40:01 |
| FLNA-02 | BANK1--PPP3CA | SLFHFLQV | 112.77 | A02:01 |
| FLNA-04 | CTDSP1--DNM3OS | VVHQVLHTR | 108.25 | A68:01 |
| LYM033 | DUSP28--PASK | QLLELEAWQL | 56.04 | A02:01 |
| LYM1322 | CEP350--RC3H1 | LLLRLQQEKV | 352.52 | A02:01 |
| LYM677 | BANK1--PPP3CA | GPAPPAVPF | 26.3 | B07:02 |
| LYM677 | CPSF6--CHMP1A | ESANIVIVRR | 9.57 | A68:01 |
| LYM677 | DNAJC10--ANKRD44 | FYERAKPPL | 123.91 | C07:02 |

#### **Supplementary Table 11: WashU vaccine candidates selected for clinical trial**

See Supp Table11.xlsx

#### **Supplementary Table 12: WashU vaccine candidates successfully manufactured for clinical trial**

See Supp Table12.xlsx

#### **Supplementary Table 13: Clinical and metadata characteristics for all patients**

See Supp Table13.xlsx

**References**

1.	Krysiak K, Gomez F, White BS, et al. Recurrent somatic mutations affecting B-cell receptor signaling pathway genes in follicular lymphoma. *Blood*. 2017;129(4):473–483.

2.	Griffith M, Griffith OL, Smith SM, et al. Genome Modeling System: A Knowledge Management Platform for Genomics. *PLoS Comput. Biol.* 2015;11(7):e1004274.

3.	[Li H. Aligning sequence reads, clone sequences and assembly contigs with BWA-MEM. *arXiv [q-bio.GN]*. 2013;](http://paperpile.com/b/a90wT9/avsy8)

4.	Picard2019toolkit. Picard toolkit. 2019;

5.	Li H. A statistical framework for SNP calling, mutation discovery, association mapping and population genetical parameter estimation from sequencing data. *Bioinformatics*. 2011;27(21):2987–2993.

6.	Barnell EK, Ronning P, Campbell KM, et al. Standard operating procedure for somatic variant refinement of sequencing data with paired tumor and normal samples. *Genet. Med.* 2019;21(4):972–981.

7.	Lonsdale J, Thomas J, Salvatore M, et al. The Genotype-Tissue Expression (GTEx) project. *Nat. Genet.* 2013;45(6):580–585.

8.	Shchetynsky K, Diaz-Gallo L-M, Folkersen L, et al. Discovery of new candidate genes for rheumatoid arthritis through integration of genetic association data with expression pathway analysis. *Arthritis Res. Ther.* 2017;19(1):19.

9.	Mo A, Marigorta UM, Arafat D, et al. Disease-specific regulation of gene expression in a comparative analysis of juvenile idiopathic arthritis and inflammatory bowel disease. *Genome Med.* 2018;10(1):48.

10.	Dvinge H, Bradley RK. Widespread intron retention diversifies most cancer transcriptomes. *Genome Med.* 2015;7(1):45.

11.	Chen R, Xia L, Tu K, et al. Longitudinal personal DNA methylome dynamics in a human with a chronic condition. *Nat. Med.* 2018;24(12):1930–1939.

12.	Madan V, Kanojia D, Li J, et al. Aberrant splicing of U12-type introns is the hallmark of ZRSR2 mutant myelodysplastic syndrome. *Nat. Commun.* 2015;6:6042.

13.	Dvinge H, Ries RE, Ilagan JO, et al. Sample processing obscures cancer-specific alterations in leukemic transcriptomes. *Proc. Natl. Acad. Sci. U. S. A.* 2014;111(47):16802–16807.

14.	Visconte V, Rogers HJ, Singh J, et al. SF3B1 haploinsufficiency leads to formation of ring sideroblasts in myelodysplastic syndromes. *Blood*. 2012;120(16):3173–3186.

15.	Ferreiro JF, Rouhigharabaei L, Urbankova H, et al. Integrative Genomic and Transcriptomic Analysis Identified Candidate Genes Implicated in the Pathogenesis of Hepatosplenic T-Cell Lymphoma. *PLoS ONE*. 2014;9(7):e102977.

16.	Hundal J, Kiwala S, McMichael J, et al. pVACtools: A Computational Toolkit to Identify and Visualize Cancer Neoantigens. *Cancer Immunol Res*. 2020;8(3):409–420.

17.	Miller CA, White BS, Dees ND, et al. SciClone: inferring clonal architecture and tracking the spatial and temporal patterns of tumor evolution. *PLoS Comput. Biol.* 2014;10(8):e1003665.

18.	Szolek A, Schubert B, Mohr C, et al. OptiType: precision HLA typing from next-generation sequencing data. *Bioinformatics*. 2014;30(23):3310–3316.

19.	Altschul SF, Gish W, Miller W, Myers EW, Lipman DJ. Basic local alignment search tool. *J. Mol. Biol.* 1990;215(3):403–410.

20.	Nielsen M, Lundegaard C, Lund O, Keşmir C. The role of the proteasome in generating cytotoxic T-cell epitopes: insights obtained from improved predictions of proteasomal cleavage. *Immunogenetics*. 2005;57(1-2):33–41.

21.	Romee R, Rosario M, Berrien-Elliott MM, et al. Cytokine-induced memory-like natural killer cells exhibit enhanced responses against myeloid leukemia. *Sci. Transl. Med.* 2016;8(357):357ra123.
