## Supplementary figures and images for "Neoantigen Landscape Supports Feasibility of Personalized Cancer Vaccine for Follicular Lymphoma"

### Supplemental Table 4

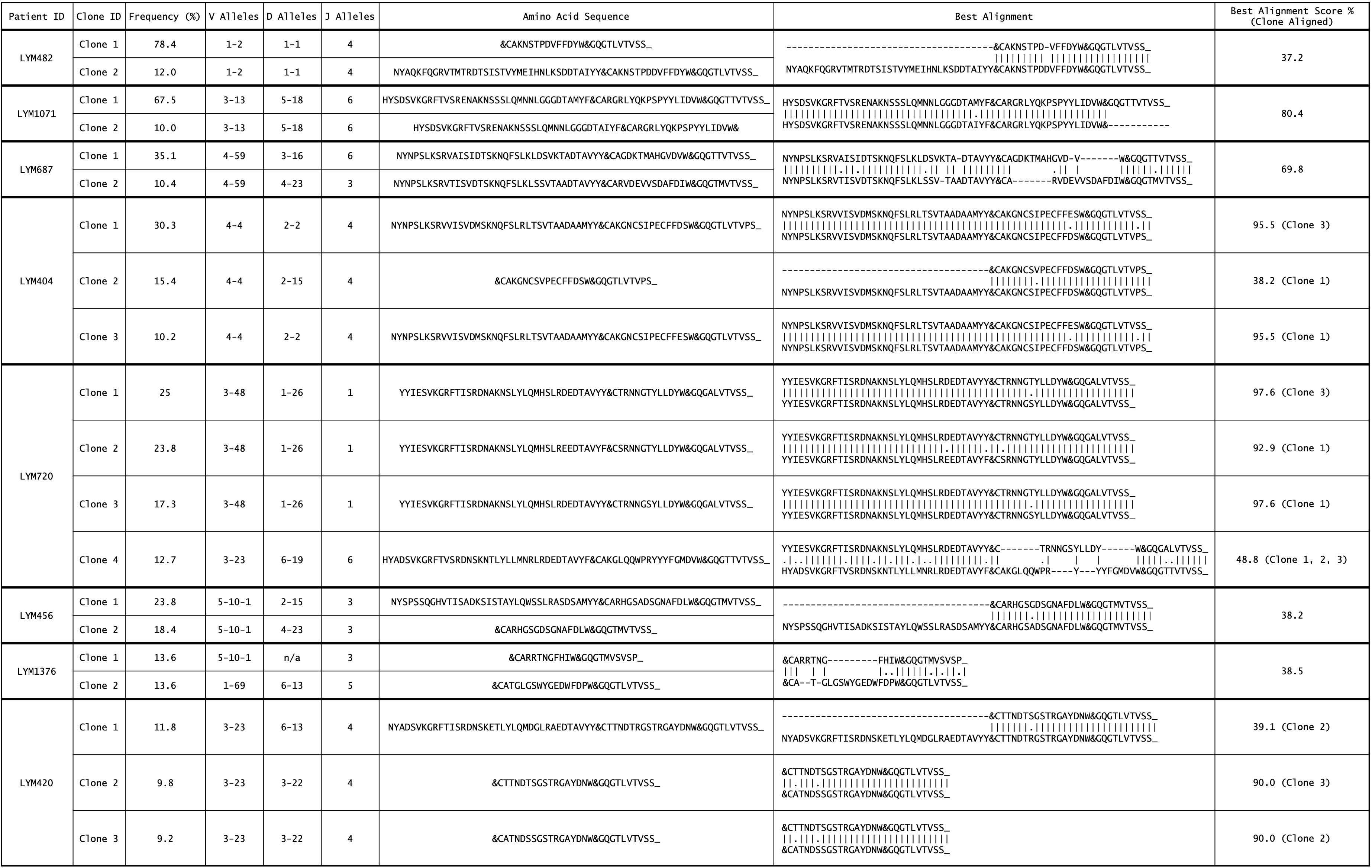

### Supplemental Table 5

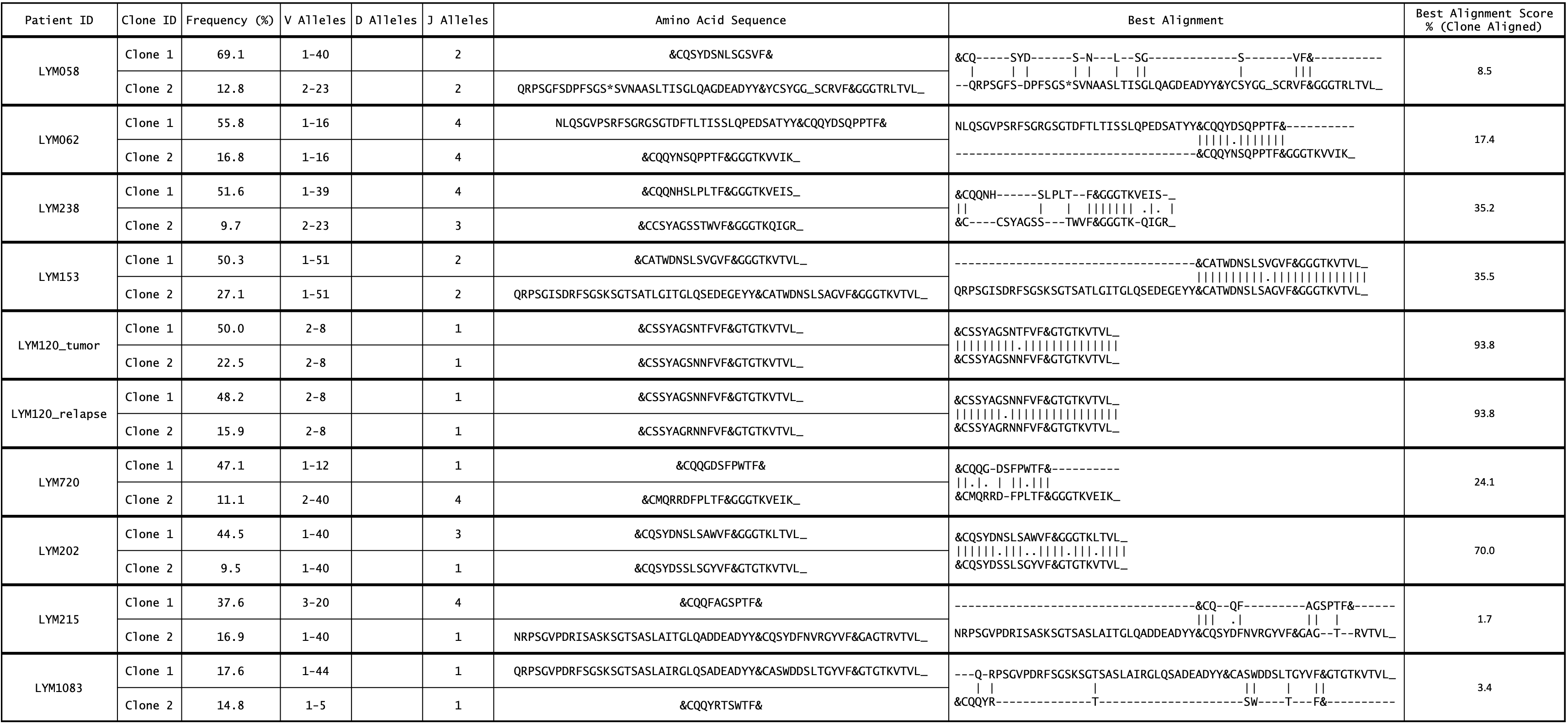
